# Costs and cost-effectiveness of influenza illness and vaccination in low- and middle-income countries: A systematic review from 2012 to 2021

**DOI:** 10.1101/2023.05.08.23289683

**Authors:** Radhika Gharpure, Anna N. Chard, Maria Cabrera Escobar, Weigong Zhou, Joseph S. Bresee, Eduardo Azziz-Baumgartner, Sarah W. Pallas, Kathryn E. Lafond

## Abstract

**Introduction:** Historically, lack of data on cost-effectiveness of influenza vaccination has been identified as a barrier to vaccine use in low- and middle-income countries. We conducted a systematic review of economic evaluations describing (1) costs of influenza illness, (2) costs of influenza vaccination programs, and (3) vaccination cost-effectiveness from low- and middle-income countries to assess if gaps persist.

**Methods:** We performed a systematic search in Medline, Embase, Cochrane Library, CINAHL, and Scopus using a combination of the following key words: “influenza” AND “cost” OR “economic.” The search included studies with publication years 2012 through 2021. We abstracted general study characteristics and data specific to each of the three areas of review.

**Results:** Of 50 included studies, 24 presented data on cost-effectiveness, 23 on cost-of-illness, and four on program costs. Represented countries were classified as upper-middle income (UMIC; n=11), lower-middle income (LMIC; n=7), and low-income (LIC; n=3). The most evaluated target groups were children (n=26 studies), older adults (n=16), and persons with chronic medical conditions (n=12); fewer studies evaluated pregnant persons (n=8), healthcare workers (n=4), and persons in congregate living settings (n=1). Costs-of-illness were generally higher in UMICs than in LMICs/LICs; however, the highest total costs, as a percent of gross domestic product and national health expenditure, were reported from an LIC. Among studies that evaluated the cost-effectiveness of influenza vaccine introduction, most (83%) interpreted at least one scenario per target group as either cost-effective or cost-saving, based on thresholds designated in the study.

**Conclusions:** Continued evaluation of the economic burden of influenza illness and costs and cost-effectiveness of influenza vaccination, particularly in low-income countries and among underrepresented target groups (e.g., healthcare workers and pregnant persons), is needed; use of standardized methodology could facilitate pooling across settings. Robust, global economic data are critical to design and maintain sustainable influenza vaccination programs.

**Summary box:** *What is already known on this topic:* Prior systematic reviews and surveys have demonstrated a need for economic data on influenza vaccination from low- and middle-income countries to inform program implementation and expansion. Standardized tools and guidance have become available in recent years to guide economic evaluations for influenza illness and vaccination in low-and middle-income countries.

*What this study adds:* This article summarizes the literature on costs of influenza illness, costs of influenza vaccination programs, and vaccination cost-effectiveness from low- and middle-income country settings during 2012–2021.

*How this study might affect research, practice, or policy:* The findings suggest value-for-money for influenza vaccination and increased interest in economic evaluations in recent years, but continued, standardized evaluation of costs and cost-effectiveness is needed, particularly from low-income countries and for underrepresented target groups.

## Introduction

Seasonal influenza vaccination is a key intervention to prevent morbidity and mortality from influenza virus infections [1]. The World Health Organization (WHO) Strategic Advisory Group of Experts on Immunization (SAGE) recommends that countries starting or expanding influenza vaccination programs prioritize specific target groups at high risk for transmission or severe disease, including healthcare workers, individuals with chronic medical conditions, older adults, and pregnant persons. Additionally, depending on priorities, available resources, and feasibility, countries might consider additional target groups for vaccination, including young children, persons in congregate living settings, disadvantaged populations, and indigenous populations [1]. As of 2018, 118 of 194 (61%) WHO member states had an influenza vaccination policy [2]; nevertheless, while low- and middle-income countries represent 40% of the world’s population and have a high burden of influenza illness [3-5], they constituted 85% of countries without a policy [2].

A 2019 survey indicated that lack of data on cost-effectiveness of influenza vaccination programs was a key barrier to initiating and expanding influenza vaccination programs in low- and middle-income countries [6]. Cost-effectiveness analyses and other economic evaluations can provide important information to guide evidence-based decision making, resource allocation, and long-term investment in vaccination by demonstrating value-for-money; however, these evaluations require accurate input data, including the costs of influenza illness, costs of vaccination, and impact of the vaccination program, in order to yield relevant and reliable results [7].

To help countries better assess the value of influenza vaccination, WHO and partners have developed standardized tools and updated guidance in recent years for economic evaluations regarding influenza illness and vaccination. These include 2016 guidance on estimating influenza economic burden [8, 9], 2016 guidance on economic evaluations for influenza vaccination, including cost-effectiveness analyses [10, 11], and a 2020 update to the Seasonal Influenza Immunization Costing Tool (SIICT) [12]. While previous systematic reviews have described economic data for influenza from low- and middle-income countries [13-17], these were generally conducted prior to the availability of these tools; more recent reviews have described data from high-income settings [16, 18, 19], focused on specific target groups [20-22], or addressed questions such as the comparative cost-effectiveness of quadrivalent and trivalent vaccines [23]. To summarize recent data and assess remaining gaps, we conducted an updated systematic review of studies describing the costs of influenza illness, costs of influenza vaccination programs, and influenza vaccination cost-effectiveness from low- and middle-income country settings within the last 10 years (2012–2021).

## Methods

This review followed the Preferred Reporting Items for Systematic Review and Meta-analysis (PRISMA) guidelines for systematic reviews and was registered at PROSPERO (international prospective register of systematic reviews) under protocol number CRD42022304803.

### Search strategy and study selection

We performed a systematic search using Medline, Embase, Cochrane Library, CINAHL, and Scopus in January 2022. The search included studies with a publication year of 2012 through 2021. Search terms were a combination of the following key words: “influenza” AND “cost” OR “economic;” specific search syntax for each database is provided in **Supplemental Table S1**.

Studies were eligible if they met the following inclusion criteria: (1) presented original, peer-reviewed findings on at least one of the following: (a) cost of illness, (b) cost of vaccination program, or (c) cost-effectiveness, cost-utility, or cost-benefit of vaccination (hereafter referred to as “cost-effectiveness studies”) for seasonal influenza; and (2) included data for at least one low- or middle-income country based on World Bank income group classification during the study period [24]. We excluded studies that: (1) did not present original or peer-reviewed findings (e.g., literature reviews, conference abstracts, and editorials); (2) only presented data about infection with or vaccination for pandemic or novel influenza viruses (e.g., influenza A(H1N1)2009 pandemic strain); or (3) included data from mid-2009 through mid-2010 that could not be disaggregated from other results, as these months were considered to represent the global influenza A(H1N1)2009 pandemic period [25].

Specifically, cost-of-illness studies were required to use a case definition of laboratory-confirmed influenza (LCI) or syndromic definitions of influenza-like-illness (ILI) and/or severe acute respiratory infection (SARI), though estimates could then be extrapolated to include other disease presentations (e.g., non-medically attended illnesses). Program cost studies were required to present population-level estimates of vaccination program costs, i.e., studies that described only cost of vaccination to the individual were excluded. Cost-effectiveness studies were required to include a comparison of influenza vaccination versus no vaccination or modifications to current vaccination program (e.g., increase in vaccination coverage); studies that only compared the cost-effectiveness of different influenza vaccine products (e.g., quadrivalent versus trivalent, adjuvanted versus non-adjuvanted, or live attenuated versus inactivated) were not included.

Titles and abstracts were independently screened by 2 reviewers (RG, AC, MC, or WZ) for eligibility, with a third reviewer resolving any conflicting decisions. English-language full texts were again reviewed by 2 reviewers (RG, AC, MC, or WZ) for eligibility, with a third reviewer resolving any conflicting decisions. Publications in other languages (Mandarin Chinese, Russian, Spanish, and Bulgarian) were reviewed by a single native-language speaker. All screening procedures were performed using Covidence, a web-based collaboration software platform for systematic reviews [26]. We also reviewed references from included studies to identify additional relevant literature for inclusion.

### Data extraction and quality assessment

Data from English-language publications were independently extracted by two reviewers (RG, AC, MC, or WZ), and disagreement was resolved by a discussion between the reviewers and consultation with a third reviewer if necessary. Data from publications in Mandarin Chinese were abstracted by a single native-language speaker (WZ); no other non-English publications met inclusion criteria.

A standardized Microsoft Excel-based data extraction form was developed to include the following information for all studies: country, study period, study methods, SAGE target group(s) represented, economic evaluation perspective, and funding source. Additionally, for cost-of-illness studies, we abstracted direct and indirect costs of all illnesses, outpatient visits, and hospitalizations. For program cost studies, we abstracted financial and economic costs both including and excluding vaccine procurement. Financial costs were incremental monetary expenditures made for the influenza vaccination program; economic costs included all financial costs as well as the value of existing resources and donations (as categorized by study authors). For cost-effectiveness studies, we abstracted the study intervention(s), comparator(s), incremental cost-effectiveness ratio (ICER), ICER interpretation, and cost-effectiveness threshold. If reported, we preferentially abstracted median values for economic variables; if medians were not reported, we abstracted mean values or ranges. We did not contact study authors to request additional unpublished data.

We used World Bank data to classify the income group of countries during the study period [24]; if countries changed income group classification during the study period, the higher classification was used. Additionally, we used World Bank data to obtain the gross domestic product (GDP) of countries during the study period [24]; for multi-year studies, the final year of the study period was used. For cost-of-illness and program cost studies, we also used World Health Organization data to obtain the Current Health Expenditure (CHE) and Domestic General Government Health Expenditure, respectively, of countries during the study period [27]. If no study period was specified, we used 3 years prior to the publication year for all relevant inputs as in prior systematic reviews [23].

For each English-language publication, two reviewers (RG, AC, MC, or WZ) assessed study quality and risk of bias using the Consolidated Health Economic Evaluation Reporting Standard (CHEERS) checklist [28]; for non-English publications, one native speaker (WZ) completed the CHEERS checklist. The checklist includes 24 criteria developed to ensure standardized reporting across economic studies; all 24 were assessed for cost-effectiveness studies, and modified sets of 13 and 15 criteria were used for cost-of-illness and program cost studies, respectively (**Supplemental Table S2**).

### Data conversion and analysis

We converted all currencies to US dollars (US$) using the International Monetary Fund official exchange rate for the nominal year [24] and then inflated all results to 2022 US$ using the U.S. Bureau of Economic Analysis GDP implicit price deflator [29, 30]. If a nominal currency year was not presented in the study, we used the final year of the study period or, if the study period was not stated, 3 years prior to the publication year. We calculated the gross national cost-of-illness and program cost, when reported, as a proportion of the national GDP and the national health expenditure. Additionally, we collated direct and indirect costs by SAGE target group and income group and reported ranges across strata. Similarly, we also collated ICER results by SAGE target group and income group and calculated the proportion of studies that interpreted findings as “cost-saving” (ICER<0), “cost-effective” (dependent on cost-effectiveness threshold specified in the study), or “not cost-effective.” All analyses were performed using SAS (version 9.4) and Microsoft Excel.

## Results

### Study characteristics and quality assessment

Of 6,614 total studies identified, 50 met eligibility criteria and were included in this review, including 43 English-language and 7 Chinese-language studies (**Figure 1, Supplemental Table S3**). Study characteristics are presented in **Table 1**; a total of 24 studies presented cost-effectiveness findings, 23 presented cost-of-illness, and four presented program costs. Studies included data from 20 country settings, which were classified as upper-middle income countries (UMICs; n=11), lower-middle income countries (LMICs; n=7), and low-income countries (LICs; n=3); one country, China, was classified as both UMIC and LMIC corresponding to multiple studies before and after an upward change in World Bank classification in 2010. These 20 countries represented 13% of 157 countries/territories classified as low- or middle-income countries in any year during 2005 (earliest year of data presented in included studies) through 2021. The most frequently evaluated SAGE target groups were children (n=26 studies, inclusive of children <18 years), older adults (n=16, inclusive of adults ≥60 years), and persons with chronic medical conditions (n=12); fewer studies evaluated pregnant persons (n=8), healthcare workers (n=4), and persons in congregate living settings (n=1).

**Figure 1:**
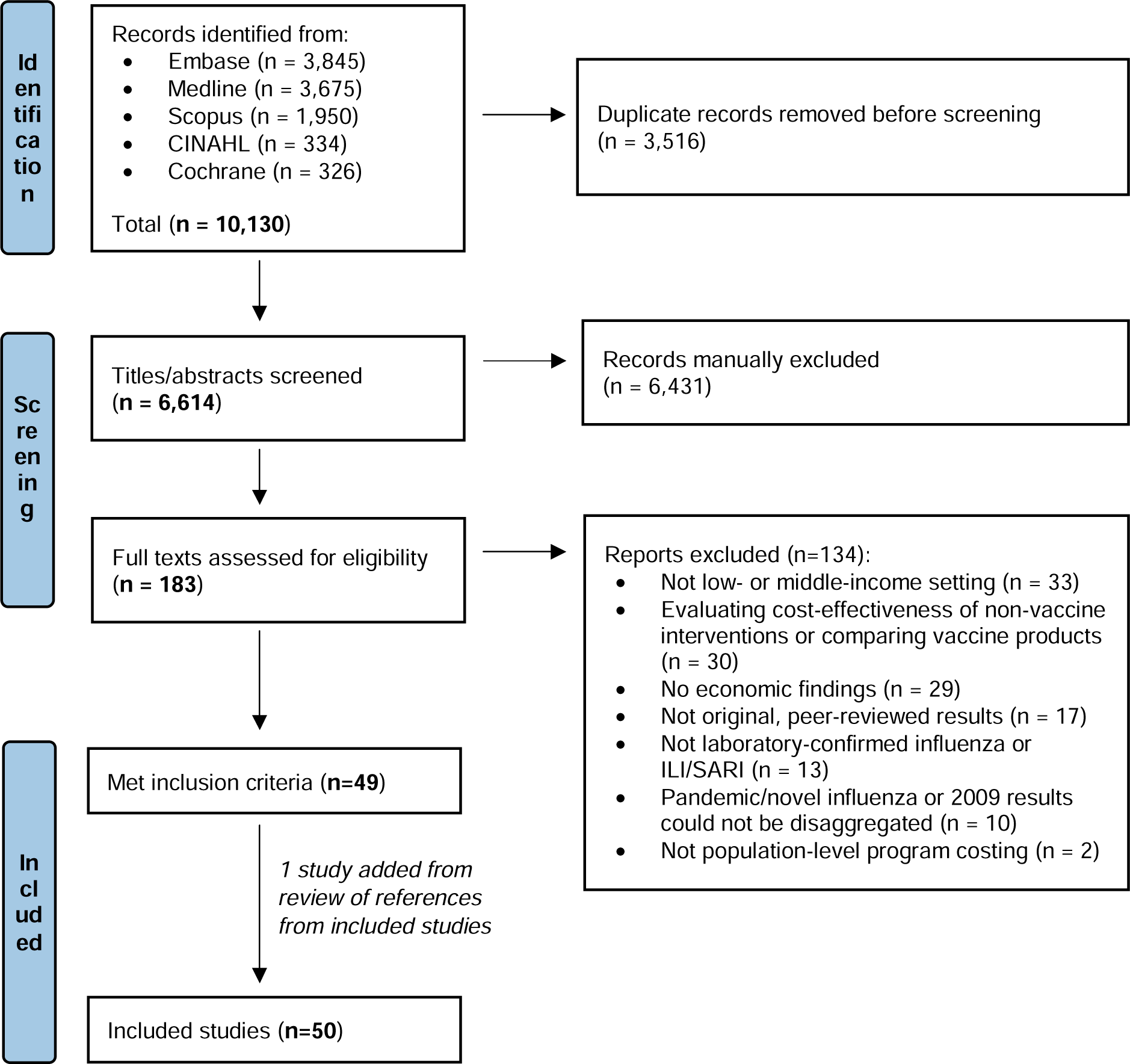
PRISMA flow diagram of study selection process.

**Table 1:** Number of included studies by income group classification, region, Strategic Advisory Committee of Experts on Immunization (SAGE) target group, and study type.

| Income group and region | Total no. studies included | Total no. countries represented | No. studies by SAGE target group<sup>1</sup> |  |  |  |  |  |  | No. studies by study type |  |  |
| --- | --- | --- | --- | --- | --- | --- | --- | --- | --- | --- | --- | --- |
|  |  |  | None (general population) | Children<sup>2</sup> | Older adults | Persons with chronic medical conditions | Pregnant persons | HCWs | Persons in congregate living settings | Cost-of-illness | Cost-of-program | Cost-effectiveness |
| **Total** | **50<sup>3</sup>** | **20<sup>4</sup>** | **13** | **26** | **16** | **12** | **8** | **4** | **1** | **23<sup>5</sup>** | **4** | **24<sup>5</sup>** |
| **Upper-middle income countries (UMIC)** | **40** | **11** | **7** | **22** | **13** | **11** | **5** | **2** | **1** | **17** | **3** | **20** |
| East Asia & Pacific | 24 | 2 | 3 | 14 | 9 | 7 | 2 | 1 | 1 | 11 | 2 | 11 |
| Latin America & Caribbean | 7 | 4 | 2 | 6 | 0 | 1 | 0 | 0 | 0 | 2 | 0 | 5 |
| Sub-Saharan Africa | 5 | 1 | 2 | 2 | 2 | 2 | 3 | 0 | 0 | 2 | 0 | 3 |
| Europe & Central Asia | 4 | 4 | 0 | 0 | 2 | 1 | 0 | 1 | 0 | 2 | 1 | 1 |
| **Lower-middle income countries (LMIC)** | **11** | **7** | **5** | **4** | **3** | **1** | **1** | **2** | **0** | **7** | **0** | **4** |
| East Asia & Pacific | 5 | 3 | 4 | 1 | 2 | 0 | 1 | 1 | 0 | 3 | 0 | 2 |
| Sub-Saharan Africa | 2 | 1 | 1 | 2 | 0 | 0 | 0 | 0 | 0 | 1 | 0 | 1 |
| Europe & Central Asia | 2 | 1 | 0 | 0 | 1 | 0 | 0 | 1 | 0 | 1 | 0 | 1 |
| Latin America & Caribbean | 1 | 1 | 0 | 1 | 0 | 0 | 0 | 0 | 0 | 1 | 0 | 0 |
| South Asia | 1 | 1 | 0 | 0 | 0 | 1 | 0 | 0 | 0 | 1 | 0 | 0 |
| **Low-income countries (LIC)** | **3** | **3** | **1** | **0** | **0** | **0** | **2** | **0** | **0** | **2** | **1** | **1** |
| Sub-Saharan Africa | 2 | 2 | 0 | 0 | 0 | 0 | 2 | 0 | 0 | 1 | 1 | 1 |
| South Asia | 1 | 1 | 1 | 0 | 0 | 0 | 0 | 0 | 0 | 1 | 0 | 0 |

<sup>1</sup>Vaccination target groups as defined in WHO SAGE guidance [1]. Several studies reported data on >1 target group.
<sup>2</sup>Although SAGE recommendations specifically reference children aged <5 years [1], publications with data for children aged <18 years were included.
<sup>3</sup>Three studies [42, 43, 69] included countries from multiple income groups and/or regions.
<sup>4</sup>One country (China) changed income classification from LMIC to UMIC in 2010 and was counted in both groups corresponding to studies assessing time periods before and after this year.
<sup>5</sup>One study [53] reported original data for both cost-of-illness and cost-effectiveness.

Quality assessment scores indicated that the quality of included studies was acceptable; median scores by study type were 12 out of 13 (92%; interquartile range [IQR] 87–100%) for cost-of-illness, 14 out of 15 (93%; IQR 93–95%) for program costs, and 23 out of 24 (96%; IQR 86–100%) for cost-effectiveness studies (**Supplemental Figure S1**); only two of 50 studies (4%) scored <75%. Of 44 studies that reported a funding source, seven (16%) were supported by pharmaceutical industry and 20 (45%) by WHO or the US Centers for Disease Control and Prevention (CDC).

### Cost-of-illness studies

The cost-per-episode of influenza illness ranged widely across studies (**Figure 2**). Twenty-three studies presented data about cost-per-episode, representing six UMICs (China [31-40], Colombia [41], Kazakhstan [42], Panama [43], Romania [42], South Africa [44, 45], and Thailand [46]), five LMICs (China [based on classification during study period] [47], El Salvador [43], India [48], Kenya [49], Ukraine [42], and Vietnam [50, 51]), and two LICs (Bangladesh [52] and Mali [53]). (**Supplemental Table S4**). Among the general population, the total cost-per-episode for outpatient visits, inclusive of direct and indirect costs, ranged from $6.24–155.92 (2022 US$); the total cost-per-episode for hospitalizations ranged from $106.85–1617.14. Among SAGE target groups, total cost-per-episode of outpatient visits and hospitalizations was $25.92–198.13 and $95.15–2202.74 for children, $38.17–164.52 and $282.37– 2729.25 for older adults, $44.13–176.79 and $847.60–1578.86 for persons with chronic medical conditions, and $5.45–36.97 and $189.98–1088.92 for pregnant persons. Costs across all target groups were generally higher in UMICs than in LMICs/LICs (**Figure 2**). Nevertheless, indirect costs comprised a greater proportion of the total costs of outpatient visits compared with hospitalizations, and a greater proportion of total costs in LMICs/LICs compared with UMICs (**Supplemental Figure S2**). Details on costs abstracted from each study are described in **Supplemental Table S4**.

**Figure 2:**
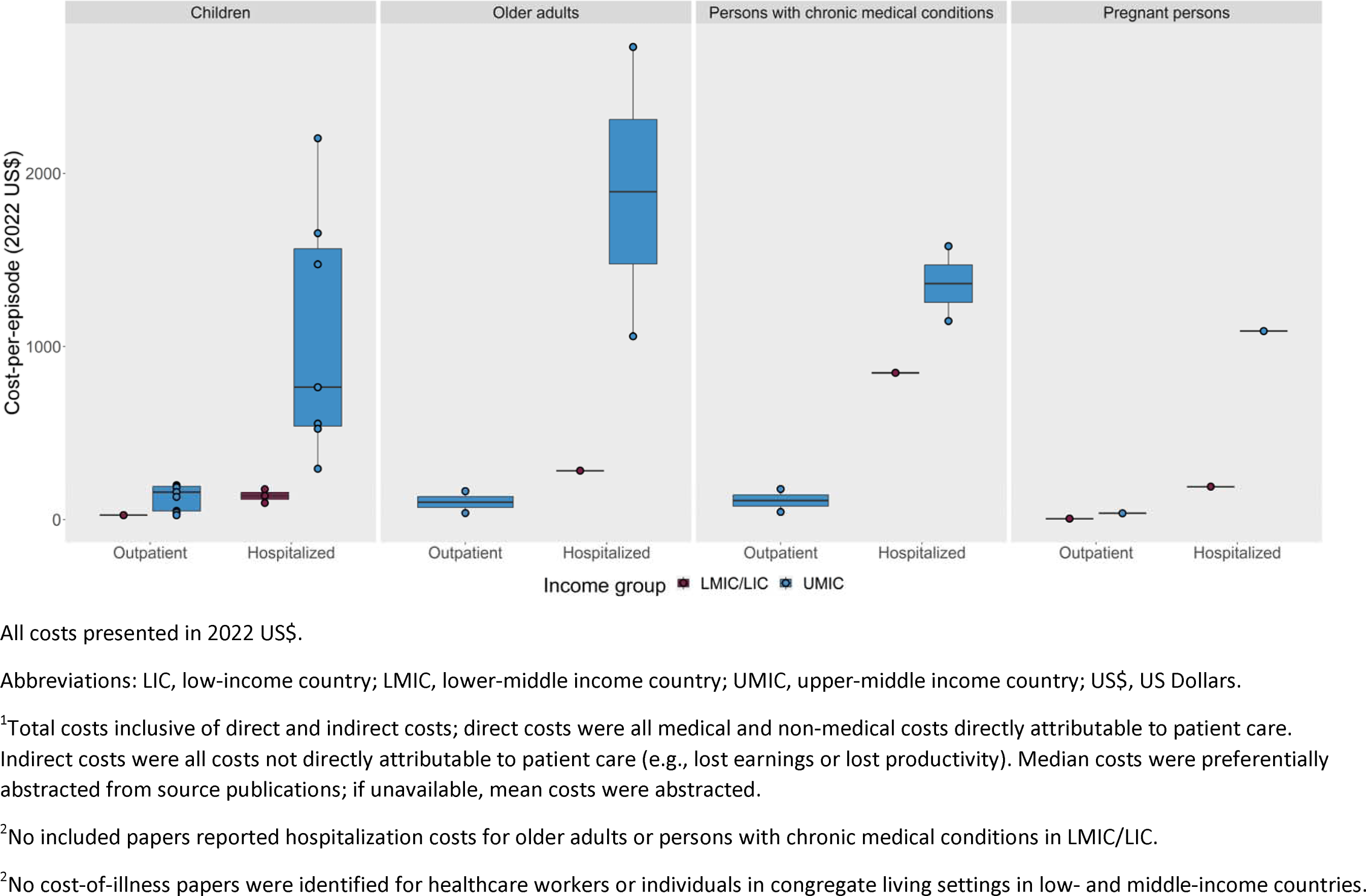
Total costs-per-episode of influenza illness, by disease severity (outpatient vs. hospitalized), income group, and Strategic Advisory Committee of Experts on Immunization (SAGE) target group, in low- and middle-income countries.

Four studies evaluated the cost-per-episode for multiple SAGE target groups [33, 38, 45, 50]. Studies from China and Vietnam found higher hospitalization costs among older adults compared with children [33, 38, 50], as well as higher costs associated with chronic medical conditions across age groups [33, 38]. In South Africa, total economic burden after incorporating rates of illness was highest for persons with chronic medical conditions, followed by children, older adults, and pregnant persons [45]. Across all studies, characteristics that impacted cost-of-illness included urbanicity (rural vs. urban) [33, 38, 47], facility type (public vs. private or level of care provision) [33, 52], and influenza season or circulating virus type [36, 39]. Only two identified studies quantified disability-adjusted life years [37] or quality-adjusted life days lost from influenza illness [35].

Seven of the 23 studies reported a total national economic burden of influenza illness for either the general population or specific SAGE target groups (**Table 2**), representing three UMICs (China [40], Romania [42], and South Africa [44, 45]), two LMICs (Kenya [49] and Ukraine [42]), and one LIC (Bangladesh [52]). Total annual costs of influenza illness in studies evaluating the general population (no specified SAGE target group) were equivalent to 0.02–0.19% of the national GDP and 0.32–7.16% of the national health expenditure; costs for any single target group were <0.01–0.02% of the national GDP and 0.01–0.42% of the national health expenditure. The highest total costs, as a percent of GDP and national health expenditure, were reported from Bangladesh [52]. Three studies accounted for non-medically attended illnesses in the estimation of national economic burden [42, 44, 45].

**Table 2:**
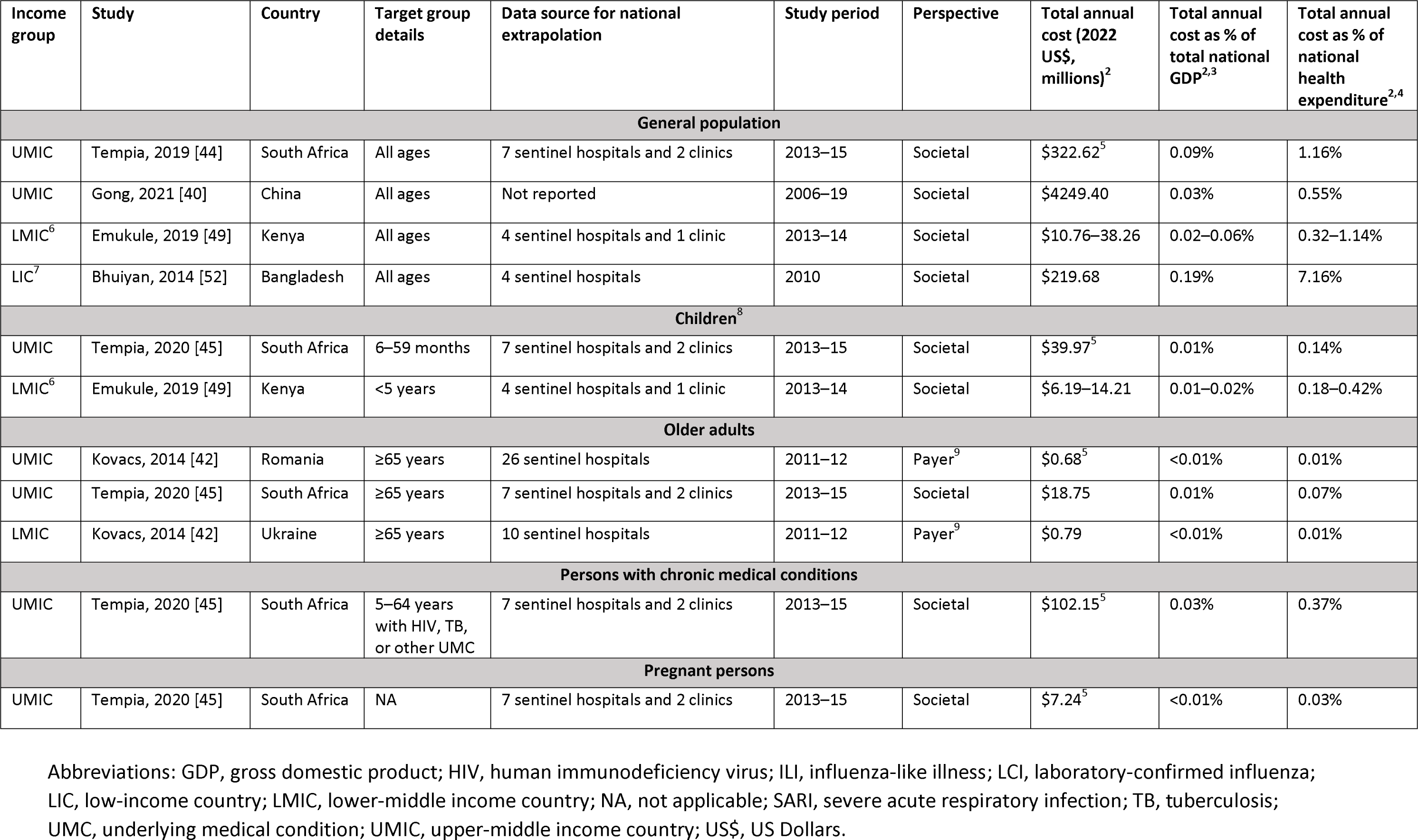
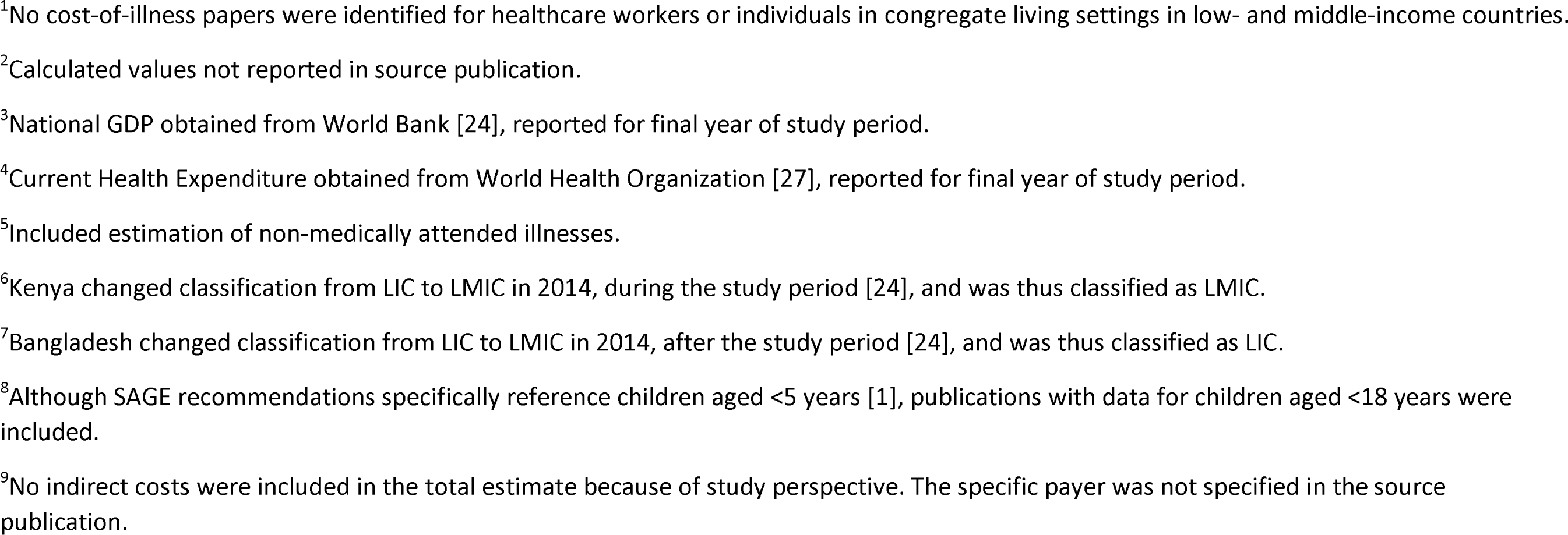
National economic burden of influenza illness, by Strategic Advisory Committee of Experts on Immunization (SAGE) target group, in low- and middle-income countries.

| Income group | Study | Country | Target group details | Data source for national extrapolation | Study period | Perspective | Total annual cost (2022 US\$, millions)<sup>2</sup> | Total annual cost as % of total national GDP<sup>2,3</sup> | Total annual cost as % of national health expenditure<sup>2,4</sup> |
| --- | --- | --- | --- | --- | --- | --- | --- | --- | --- |
| **General population** |  |  |  |  |  |  |  |  |  |
| UMIC | Tempia, 2019 [44] | South Africa | All ages | 7 sentinel hospitals and 2 clinics | 2013–15 | Societal | \$322.62<sup>5</sup> | 0.09% | 1.16% |
| UMIC | Gong, 2021 [40] | China | All ages | Not reported | 2006–19 | Societal | \$4249.40 | 0.03% | 0.55% |
| LMIC<sup>6</sup> | Emukule, 2019 [49] | Kenya | All ages | 4 sentinel hospitals and 1 clinic | 2013–14 | Societal | \$10.76–38.26 | 0.02–0.06% | 0.32–1.14% |
| LIC<sup>7</sup> | Bhuiyan, 2014 [52] | Bangladesh | All ages | 4 sentinel hospitals | 2010 | Societal | \$219.68 | 0.19% | 7.16% |
| **Children<sup>8</sup>** |  |  |  |  |  |  |  |  |  |
| UMIC | Tempia, 2020 [45] | South Africa | 6–59 months | 7 sentinel hospitals and 2 clinics | 2013–15 | Societal | \$39.97<sup>5</sup> | 0.01% | 0.14% |
| LMIC<sup>6</sup> | Emukule, 2019 [49] | Kenya | <5 years | 4 sentinel hospitals and 1 clinic | 2013–14 | Societal | \$6.19–14.21 | 0.01–0.02% | 0.18–0.42% |
| **Older adults** |  |  |  |  |  |  |  |  |  |
| UMIC | Kovacs, 2014 [42] | Romania | ≥65 years | 26 sentinel hospitals | 2011–12 | Payer<sup>9</sup> | \$0.68<sup>5</sup> | <0.01% | 0.01% |
| UMIC | Tempia, 2020 [45] | South Africa | ≥65 years | 7 sentinel hospitals and 2 clinics | 2013–15 | Societal | \$18.75 | 0.01% | 0.07% |
| LMIC | Kovacs, 2014 [42] | Ukraine | ≥65 years | 10 sentinel hospitals | 2011–12 | Payer<sup>9</sup> | \$0.79 | <0.01% | 0.01% |
| **Persons with chronic medical conditions** |  |  |  |  |  |  |  |  |  |
| UMIC | Tempia, 2020 [45] | South Africa | 5–64 years with HIV, TB, or other UMC | 7 sentinel hospitals and 2 clinics | 2013–15 | Societal | \$102.15<sup>5</sup> | 0.03% | 0.37% |
| **Pregnant persons** |  |  |  |  |  |  |  |  |  |
| UMIC | Tempia, 2020 [45] | South Africa | NA | 7 sentinel hospitals and 2 clinics | 2013–15 | Societal | \$7.24<sup>5</sup> | <0.01% | 0.03% |

Abbreviations: GDP, gross domestic product; HIV, human immunodeficiency virus; ILI, influenza-like illness; LCI, laboratory-confirmed influenza; LIC, low-income country; LMIC, lower-middle income country; NA, not applicable; SARI, severe acute respiratory infection; TB, tuberculosis; UMC, underlying medical condition; UMIC, upper-middle income country; US\$, US Dollars.
<sup>1</sup>No cost-of-illness papers were identified for healthcare workers or individuals in congregate living settings in low- and middle-income countries.
<sup>2</sup>Calculated values not reported in source publication.
<sup>3</sup>National GDP obtained from World Bank [24], reported for final year of study period.
<sup>4</sup>Current Health Expenditure obtained from World Health Organization [27], reported for final year of study period.
<sup>5</sup>Included estimation of non-medically attended illnesses.
<sup>6</sup>Kenya changed classification from LIC to LMIC in 2014, during the study period [24], and was thus classified as LMIC.
<sup>7</sup>Bangladesh changed classification from LIC to LMIC in 2014, after the study period [24], and was thus classified as LIC.
<sup>8</sup>Although SAGE recommendations specifically reference children aged <5 years [1], publications with data for children aged <18 years were included.
<sup>9</sup>No indirect costs were included in the total estimate because of study perspective. The specific payer was not specified in the source publication.

### Program cost studies

Four studies evaluated the cost of influenza vaccination programs (**Table 3**): three with findings from UMICs (Albania [54], China [55], and Thailand [56]) and one from an LIC (Malawi [57]). Of these, two evaluated the cost of a program targeting pregnant persons [56, 57], one evaluated a program targeting healthcare workers [54], and one evaluated a program targeting multiple SAGE target groups (older adults, persons with chronic medical conditions, children <5 years, pregnant persons, and healthcare workers) [55]. Two studies used the WHO SIICT [12, 54, 57]. The total annual cost of program was equivalent to <0.01–0.04% of the national GDP and 0.06–4.78% of the national health expenditure; the highest proportion of health expenditure was reported in the study vaccinating multiple target groups. Vaccine procurement represented a large proportion of total costs (89% financial and 44% economic costs in Albania [54] and 1% financial and 82% economic costs, assuming donated vaccine, in Malawi [57]). Across studies, the total cost per dose administered ranged from $0.62–5.20 (financial) and $0.81– 13.72 (economic), inclusive of vaccine purchase or donation.

**Table 3:**
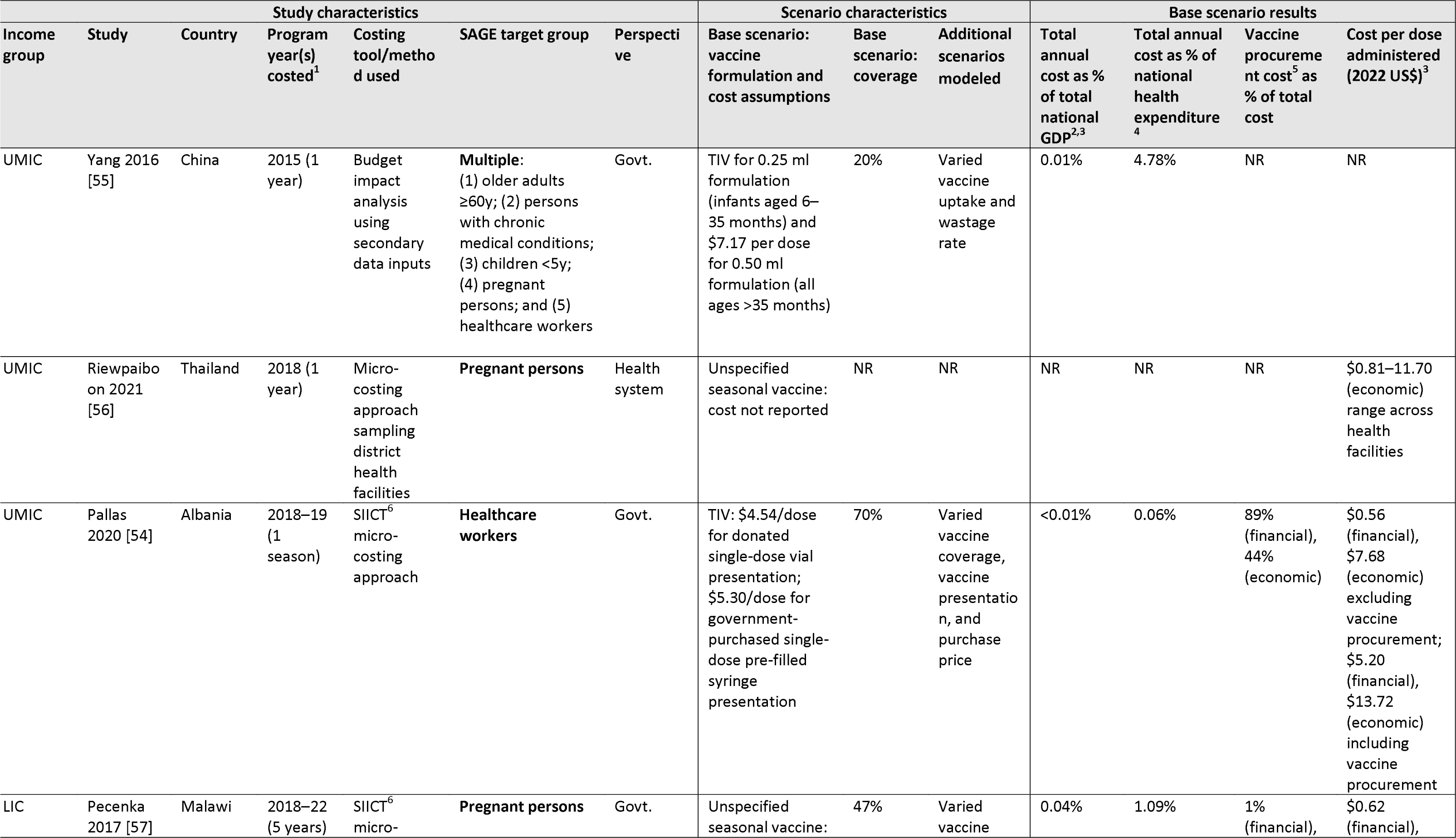
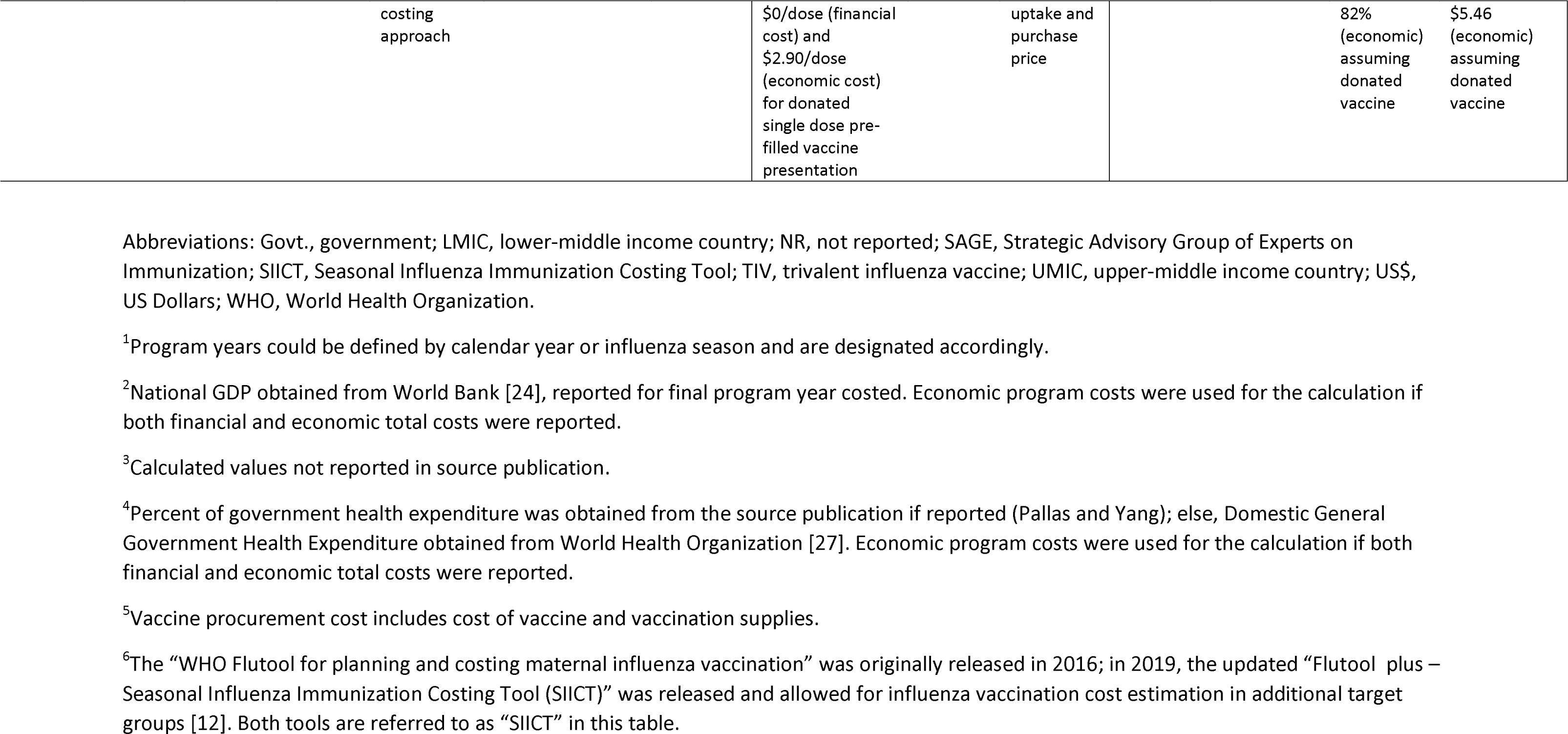
Costs of national influenza vaccination programs in low- and middle-income countries.

### Cost-effectiveness studies

Twenty-four studies presented data on cost-effectiveness of influenza vaccination (**Supplemental Table S5**), representing seven UMICs (Argentina [58], China [59-64], Colombia [65], Mexico [66-68], South Africa [69-71], Thailand [72-76], and Turkiye [77]), four LMICs (Kenya [78], Lao PDR [79], Ukraine [80], and Vietnam [69]), and one LIC (Mali [53]). Twenty (83%) studies evaluated influenza vaccine introduction (i.e., vaccination compared with no vaccination), two evaluated the effect of increased vaccination coverage on an existing program, and two evaluated combinations of new introduction and increased coverage for different target groups. Cost-effectiveness thresholds varied greatly across studies; most (n=15/24; 63%) used a threshold within one to three times the GDP per capita, three (13%) used other country-specific thresholds, one (4%) intentionally did not report a threshold, and the remaining five (21%) did not provide any details about thresholds.

Among the 20 studies that evaluated the cost-effectiveness of vaccine introduction, eight provided results for children, five for older adults, four for persons with chronic conditions, four for pregnant persons, two for healthcare workers, and one for persons in congregate living settings. Most (83%) interpreted at least one modeled scenario for each SAGE target group as either cost-effective (based on designated cost-effectiveness threshold) or cost-saving (ICER<0) (**Figure 3**). The number of studies that identified results as cost-saving were 3/8 (38%) for children, 1/5 (20%) for older adults, 2/4 (50%) for persons with chronic medical conditions, 1/4 (25%) for pregnant persons, and 2/2 (100%) for healthcare workers. Similarly, the number of studies that identified results as cost-effective were 3/8 (38%) for children, 3/5 (60%) for older adults, 2/4 (50%) for persons with chronic medical conditions, 3/4 (75%) for pregnant persons, and 1/1 (100%) for persons in congregate living settings. Only three studies interpreted all modeled scenarios for a particular target group as not cost-effective: 2/8 (25%) evaluating cost-effectiveness among children [71, 78] and 1/5 (20%) among older adults [64].

**Figure 3:**
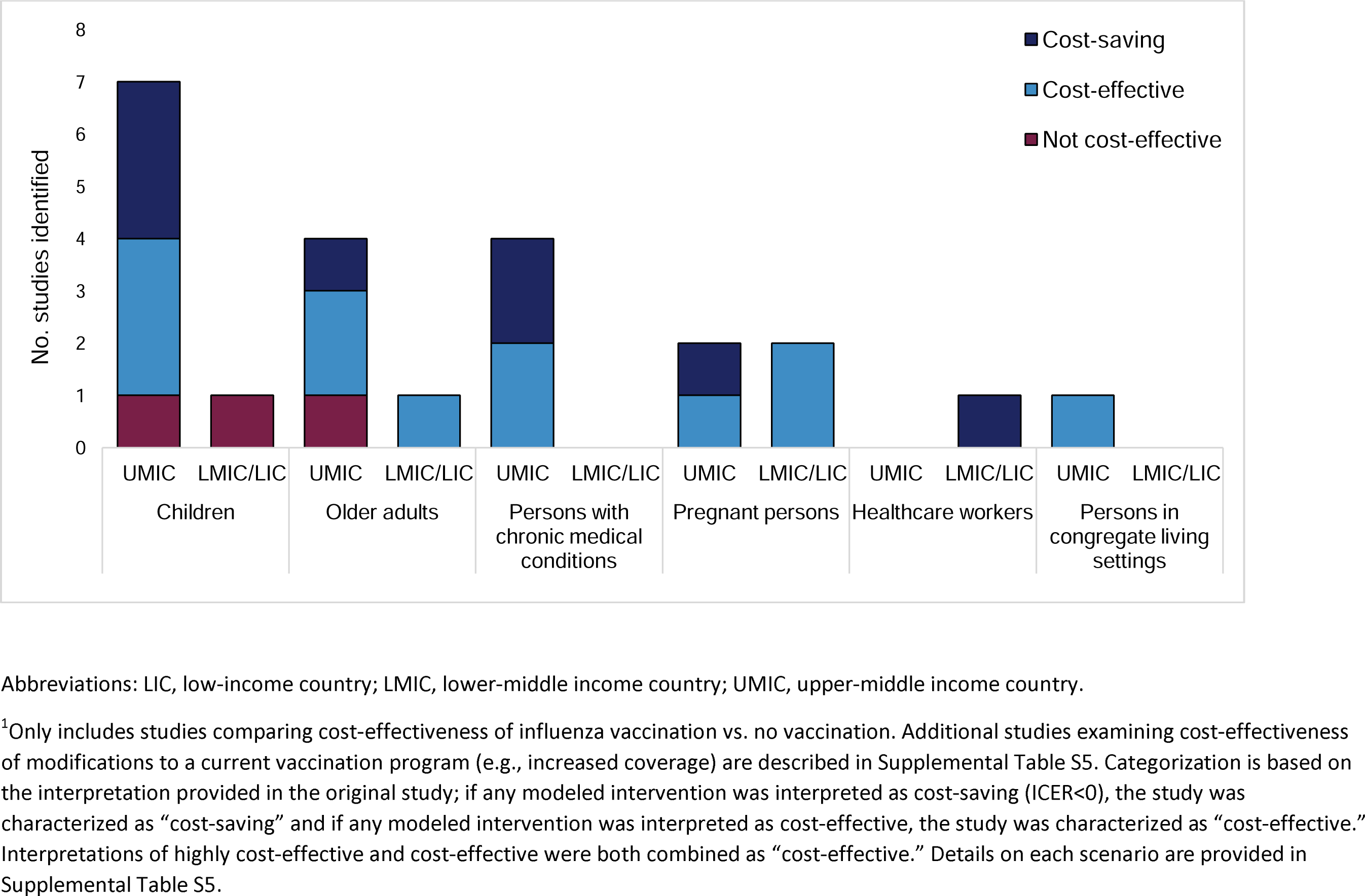
Cost-effectiveness results of studies evaluating influenza vaccination, by Strategic Advisory Committee of Experts on Immunization (SAGE) target group, in low- and middle-income countries.

Two studies assessed the cost-effectiveness of influenza vaccine introduction for multiple target groups; of these, the target groups with the greatest value-for-money were healthcare workers in Laos (cost-saving; other groups evaluated were pregnant persons and older adults, found to be cost-effective) [79], and pregnant persons and persons with chronic medical conditions in South Africa (cost saving; other groups evaluated were older adults, found to be cost-effective, and children, not found to be cost-effective) [71]. The three variables most commonly identified to influence the ICER were annual incidence of influenza (n=9) [53, 58, 59, 69-71, 74, 77, 79], vaccine effectiveness (n=9 studies) [58, 63, 65, 70-73, 77, 79], and cost of vaccine (n=6) [53, 70, 71, 73, 74, 78]; some studies demonstrated that variation in the attack rate [59, 69, 71, 74] or vaccine effectiveness [71] change the interpretation of cost-effectiveness results (cost-saving, cost-effective, or not-cost-effective).

Additionally, seven papers looked at different prioritization strategies within SAGE target groups by underlying health conditions [70, 72] or age [58, 59, 73, 75, 78]; among pregnant persons in South Africa, prioritization of people living with HIV reduced the ICER (though not statistically significant) [70], and among persons with underlying coronary heart disease in Thailand, restricting to only persons with angina reduced the ICER, whereas restricting to persons with cardiac arrest/myocardial infarction increased the ICER (no longer cost-effective) [72]. Results by age were also mixed; two studies among children found lower ICERs for vaccinating younger children (6–23 months vs. 2–5 years or 6–14 years in Kenya [78] and 6–59 months vs. 5–14 years in China [59]), two studies among children found lower ICERs for vaccinating wider age ranges (6 months–5 years vs. 6–23 months or 6–36 months in Argentina [58]) or older children (12–17 years vs. 2–5 years or 6–11 years in Thailand [73]), and a study among persons with underlying heart disease in Thailand found a lower ICER for persons aged ≥50 years compared with ≥40 years or ≥60 years [75].

## Discussion

The 50 studies identified in this review suggest an increased momentum to generate economic evidence about influenza illness and vaccination from low- and middle-income countries during 2012 – 2021; a previous review using a similar search strategy identified only 22 cost-of-illness or cost-effectiveness studies from low- and middle-income countries [15], and another identified nine cost-effectiveness/cost-benefit/cost-utility studies [13], both prior to 2012. The release of updated tools and guidance by WHO, as well as technical and financial support by WHO, CDC, and other international partners, have facilitated this expansion of the evidence base, emphasizing the utility of global and multinational collaborations in strengthening influenza vaccination programs worldwide. Recent additions to the literature include studies from LMIC/LICs, studies representing Sub-Saharan Africa, South Asia, and middle-income European countries, and studies focused on pregnant persons; none of these were represented in the previous reviews, which only identified data from UMICs in East Asia, Latin America, and Europe [13, 15]. However, disparities remain by income group and region; LICs are still very underrepresented, and no studies from low- and middle-income countries in the Middle East and North Africa region were identified in our review.

Additionally, pregnant persons, healthcare workers, and persons in congregate living settings remain especially underrepresented in economic evaluations. Healthcare workers are of particular interest because of the potential benefit of vaccination to themselves and the greater health system [81]; in 2018, Gavi, the Vaccine Alliance, expressed interest in assessing the feasibility and impact of routine influenza immunization of healthcare workers to support epidemic and pandemic influenza preparedness [82]. Additionally, the Gavi 5.1 strategy, presented in December 2022 [83], included a strategic focus on enhancing outbreak and pandemic response, to which healthcare worker vaccination might contribute. To date, global literature about cost-effectiveness and other evidence for influenza vaccination among healthcare workers remains limited [81, 84], but notably, we identified both a cost-saving result for healthcare worker vaccination in Lao PDR [79] and in Ukraine [80], suggesting high value-for-money. Additional data are needed to strengthen the evidence to optimize influenza vaccination in this target group.

Among all cost-of-illness studies, we found that the cost-per-episode estimates for influenza outpatient visits and hospitalizations varied widely. Per-episode costs were generally greater in higher income settings (i.e., UMICs compared with LMICs/LICs), likely reflecting higher costs of care, but the national economic burden among the general population, which ranged from <1%–7% of the national health expenditure, was highest in an LIC (Bangladesh [52]). More studies from LICs are needed to further evaluate disparities among income groups. As recommended by WHO, many studies used data from ILI/SARI sentinel surveillance sites for estimating economic burden; these surveillance systems can serve as a valuable data source but are not typically designed to capture non-medically attended illnesses [85] or non-respiratory disease outcomes [86], thereby likely underestimating the true economic burden of influenza. In South Africa for example, estimates obtained among patients meeting a SARI/ILI case definition underestimated the total economic burden by approximately 65% [44]; thus, comprehensive strategies and innovative strategies are needed to better characterize economic burden. Finally, characteristics of the underlying population in a particular setting, such as age structure and prevalence of underlying medical conditions, might affect costs across target groups; for example, in South Africa, where the highest burden was in individuals with chronic medical conditions, this was impacted by HIV and TB prevalence in the population [44, 45]. Thus, economic evaluations that address multiple target groups in a particular setting, rather than a single target group, can provide valuable evidence to inform local vaccination policy; given limited resources for vaccination programs, such comparisons could assist with target group prioritization.

We identified only four program cost studies within the last 10 years, indicating a need for more evaluations in low- and middle-income countries. Vaccine delivery cost studies can provide direct evidence to policymakers to make decisions on vaccine introduction, plan budgets and financing strategies for rollout, and identify efficiencies in service delivery [87]. In fact, the WHO SIICT [12] and other costing methods can be used even in the absence of an existing program, as performed in Malawi [57]. The four studies that we identified indicated that influenza vaccination programs generally cost a small fraction compared to the national GDP (≤0.04% in these studies) or national health expenditure (≤1% per each individual target group covered in these studies). Vaccine procurement was a major driver of program costs in both studies that disaggregated this component, representing 82% of the economic costs (including the value of donated resources) of the hypothetical maternal vaccination program using donated vaccine in Malawi [57], and 89% of the financial and 44% of the economic costs of a healthcare worker vaccination program utilizing a combination of government-procured and donated vaccines in Albania [54]. This underscores the importance of sustainable financing and procurement strategies to support access to influenza vaccines and enable successful program implementation, consistent with lessons learned from other vaccine introductions [88]. Again, as costs may vary across target groups, evaluating program costs in multiple groups within a given country context could provide useful data for resource prioritization.

Among cost-effectiveness studies identified in this review, most reported at least one cost-saving or cost-effective vaccination scenario per target group assessed; however, results were significantly impacted by variables such as influenza incidence, vaccine effectiveness, cost of vaccine, and vaccine coverage, as well as by prioritization within target groups (e.g., by age or specific underlying health conditions). Strategies to address this variability include use of at least 5 years of data to assess disease burden, if available, and use of sensitivity analyses among ranges of plausible values, for example including vaccine effectiveness estimates from years with high and low vaccine match [11]. Future studies could use innovative approaches to more completely characterize the total disease and economic burden of influenza, as well as additional endpoints for vaccine effectiveness (illness attenuation) and indirect protection from vaccination [7]. Finally, the use of appropriate cost-effectiveness thresholds in low- and middle-income settings warrants further discussion [89, 90]. In our review, among only three studies that did not identify any cost-effective scenarios, two used a cost-effectiveness threshold less than GDP per capita [71, 78]; however, using 1–3 times GDP per capita would have resulted in a cost-effective result in both. Use of context-specific thresholds reflecting local preferences [91], such as local health opportunity costs [92], might provide more valuable information to guide investment decisions than thresholds of 1–3 times GDP per capita [89, 90, 93, 94].

This review is subject to several notable limitations. First, the inclusion/exclusion criteria used (e.g., estimates derived from LCI or ILI/SARI case definition; no comparison of vaccine formulations) undercount the total number of economic studies from low- and middle-income countries within the past 10 years. Multiple other studies have evaluated costs of acute respiratory illness, of which influenza is an important etiology, or addressed other economic questions, such as the cost-effectiveness of quadrivalent vs. trivalent vaccine [23, 95], and were not captured here. Influenza illness might also present as non-respiratory outcomes [86], and thus the economic burden of influenza is underestimated in most studies that restrict to syndromic surveillance for ILI/SARI [85]. Second, target group definitions vary across countries, with variation in age cut-offs for children and older adults and prioritization of specific chronic medical conditions, but all results per target group were summarized together in this review due to the small numbers of publications, potentially missing nuances of within-group differences. Relatedly, although SAGE recommendations specifically reference children aged <5 years [1], all publications with data for children aged <18 years were included. Finally, we found substantial heterogeneity in the methodology and data inputs used across studies; in fact, as discussed previously, influenza itself intrinsically varies in annual incidence, disease severity, and vaccine effectiveness across seasons. As previously discussed, we did not conduct meta-analyses because of this variability, though methods for meta-analysis of economic data are available [96] and have been used in other reviews that focus predominantly on high-income settings [21].

This review also uncovered opportunities to provide evidence on policy-relevant questions that currently have limited evidence. First, we did not identify any studies taking an employer payer’s perspective; however, studies utilizing this approach could provide valuable policy-relevant information to encourage vaccination among employees or to encourage employer-supported vaccination programs [97] as a pathway to broader influenza vaccine availability. Second, few studies quantified preference-based outcomes such as disability-adjusted life years [40] or quality-adjusted life days from influenza illness [38]; continued efforts for valuation of these outcomes could inform context-specific inputs for future cost-effectiveness analyses. Third, we only identified one included study that evaluated cost-effectiveness of influenza vaccination coadministered with another vaccine (pneumococcal vaccine) [65]; a few additional studies addressing coadministration were excluded because they did not provide results for influenza vaccination alone. Given opportunities to coadminister influenza vaccine with other vaccines across the life course, including COVID-19 vaccine [98], evaluation of shared costs in program cost or cost-effectiveness studies might incentivize integrated vaccine implementation. Fourth, we found only two studies that considered non-respiratory disease outcomes (cardiovascular disease events) in cost-effectiveness analyses, both among persons with underlying heart disease in Thailand [72, 75]; as previously discussed, inclusion of non-respiratory disease outcomes could better characterize the full impact of influenza vaccination [86]. Similarly, innovative strategies might address the broader impact of vaccines, such as impact on childhood development, household behavior, economic growth, political stability, and health equity [99, 100]

## Conclusions

Continued evaluation of costs and cost-effectiveness is useful to drive evidence-based vaccine policy development, implementation, and refinement and global investment in influenza vaccination. Additional studies from low-income countries and underrepresented target groups (e.g., pregnant persons, healthcare workers, and persons in congregate living settings) would strengthen the evidence of value-for-money. Standardization of research agenda [1] and methodology across future evaluations, including considerations to capture the full spectrum of influenza-associated illness, could allow for pooled estimates and meta-analyses. Global, regional, and country-specific data on the economics of vaccination, including costs of vaccination programs, costs of avertable illnesses, and cost-effectiveness, are instrumental for policymaking, resource allocation, and investment for expanded and sustainable influenza vaccination programs.

## Data Availability

All data produced in the present work are contained in the manuscript.

## Acknowledgements

We thank Rakhat Akmatova, Silvia Bino, and Tat Yao for screening of non-English publications; Britni Burkhardsmeier for administrative project support; Joanna Taliano for search strategy support; and Chris Chadwick, Stefano Tempia, and members of the Partnership for Influenza Vaccine Introduction (PIVI) technical working group for their insightful review and suggestions.

## Disclaimer

The findings and conclusions in this report are those of the authors and do not necessarily represent the official position of the US Centers for Disease Control and Prevention.

## Supplementary materials

**Table S1:** Search terms for systematic review, by database.

| Database | Strategy | Run Date |
| --- | --- | --- |
| **Medline**<br>**(OVID)**<br>**1946-** | (influenza* OR flu OR H1N1)<br>AND<br>Cost* OR economic* OR ec.fs<br>AND<br>Exp Animals/ NOT exp humans/<br>2012 - 2021 | 01/31/2022 |
| **Embase**<br>**(OVID)**<br>**1974-** | Exp Influenza/ OR (influenza* OR flu OR H1N1).ti,ab.<br>AND<br>Cost* OR economic*<br>AND<br>Exp Animal/ NOT exp human/<br>NOT<br>Conference abstract.pt<br>2012 – 2021; not Pubmed/Medline | 01/31/2022 |
| **Cochrane Library** | (influenza* OR flu OR H1N1):ti,ab<br>AND<br>(Cost* OR economic*):ti,ab<br>2012 – 2021 | 01/31/2022 |
| **CINAHL**<br>**(EbscoHost)** | (TI (influenza* OR flu OR H1N1)) OR (AB (influenza* OR flu OR H1N1))<br>AND<br>(TI (Cost* OR economic*)) OR (AB (Cost* OR economic*))<br>2012 – 2021; exclude Medline records | 01/31/2022 |
| **Scopus** | TITLE-ABS-KEY(influenza* OR flu OR H1N1) AND TITLE-ABS-KEY(Cost* OR economic*) AND NOT INDEX(medline)<br>2012 – 2021 | 01/31/2022 |

**Table S2:** Modified Consolidated Health Economic Evaluation Reporting Standard (CHEERS) criteria^1^ used for quality assessment.

| CHEERS criterion | Description | Evaluated for cost-of-illness studies | Evaluated for cost-of-program studies | Evaluated for cost-effectiveness studies |
| --- | --- | --- | --- | --- |
| Title | Identify the study as an economic evaluation or use more specific terms such as “cost-effectiveness analysis”, and describe the interventions compared. | ? | ? | ? |
| Abstract | Provide a structured summary of objectives, perspective, setting, methods (including study design and inputs), results (including base case and uncertainty analyses), and conclusions. | ? | ? | ? |
| Background and objectives | Provide an explicit statement of the broader context for the study. Present the study question and its relevance for health policy or practice decisions. | ? | ? | ? |
| Target population and subgroups | Describe characteristics of the base case population and subgroups analyzed, including why they were chosen. | ? | ? | ? |
| Setting and location | State relevant aspects of the system(s) in which the decision(s) need(s) to be made. | ? | ? | ? |
| Study perspective | Describe the perspective of the study and relate this to the costs being evaluated. | ? | ? | ? |
| Comparators | Describe the interventions or strategies being compared and state why they were chosen. |  |  | ? |
| Time horizon | State the time horizon(s) over which costs and consequences are being evaluated and say why appropriate. |  | ? | ? |
| Discount rate | Report the choice of discount rate(s) used for costs and outcomes and say why appropriate. |  | ? | ? |
| Choice of health outcomes | Describe what outcomes were used as the measure(s) of benefit in the evaluation and their relevance for the type of analysis performed. |  |  | ? |
| Measurement of effectiveness | Describe fully the design features of the single effectiveness study and why the single study was a sufficient source of clinical effectiveness data. **OR**<br>Describe fully the methods used for identification of included studies and synthesis of clinical effectiveness data. |  |  | ? |
| Measurement and valuation of preference-based outcomes | If applicable, describe the population and methods used to elicit preferences for outcomes |  |  | ?<br>(if QALY or DALY results presented) |
| Estimating resources and costs | Describe approaches used to estimate resource use associated with the alternative interventions. Describe primary or secondary research methods for valuing each resource item in terms of its unit cost. Describe any adjustments made to approximate to opportunity costs. **OR**<br>Describe approaches and data sources used to estimate resource use associated with model health states. Describe primary or secondary research methods for valuing each resource item in terms of its unit cost. Describe any adjustments made to approximate to opportunity costs. | ? | ? | ? |
| Currency, price date, and conversion | Report the dates of the estimated resource quantities and unit costs. Describe methods for adjusting estimated unit costs to the year of reported costs if necessary. Describe methods for converting costs into a common currency base and the exchange rate. | ? | ? | ? |
| Choice of model | Describe and give reasons for the specific type of decision analytical model used. Providing a figure to show model structure is strongly recommended. |  |  | ? |
| Assumptions | Describe all structural or other assumptions underpinning the decision-analytical model. |  |  | ? |
| Analytical methods | Describe all analytical methods supporting the evaluation. This could include methods for dealing with skewed, missing, or censored data; extrapolation methods; methods for pooling data; approaches to validate or make adjustments (such as half cycle corrections) to a model; and methods for handling population heterogeneity and uncertainty. | ? | ? | ? |
| Study parameters | Report the values, ranges, references, and, if used, probability distributions for all parameters. Report reasons or sources for distributions used to represent uncertainty where appropriate. Providing a table to show the input values is strongly recommended. | ? | ? | ? |
| Incremental costs and outcomes | For each intervention, report mean values for the main categories of estimated costs and outcomes of interest, as well as mean differences between the comparator groups. If applicable, report incremental cost-effectiveness ratios. |  |  | ? |
| Characterizing uncertainty | Describe the effects of sampling uncertainty for the estimated incremental cost and incremental effectiveness parameters, together with the impact of methodological assumptions (such as discount rate, study perspective). **OR**<br>Describe the effects on the results of uncertainty for all input parameters, and uncertainty related to the structure of the model and assumptions. |  |  | ? |
| Characterizing heterogeneity | If applicable, report differences in costs, outcomes, or cost-effectiveness that can be explained by variations between subgroups of patients with different baseline characteristics or other observed variability in effects that are not reducible by more information. |  |  | ? |
| Study findings, limitations, generalizability, and current knowledge | Summarize key study findings and describe how they support the conclusions reached. Discuss limitations and the generalizability of the findings and how the findings fit with current knowledge. | ? | ? | ? |
| Source of funding | Describe how the study was funded and the role of the funder in the identification, design, conduct, and reporting of the analysis. Describe other non-monetary sources of support. | ? | ? | ? |
| Conflicts of interest | Describe any potential for conflict of interest of study contributors in accordance with journal policy. In the absence of a journal policy, we recommend authors comply with International Committee of Medical Journal Editors recommendations. | ? | ? | ? |
| **Total possible score** |  | **13** | **15** | **23–24** |

<sup>1</sup>The full CHEERS criteria assessment [28] was performed for cost-effectiveness studies; a modified set of relevant criteria were assessed for cost-of-illness and cost-of-program studies.

**Table S3:** Description of included studies, by country.

| Country | Author, year | Study type | SAGE target group(s) |  |  |  |  |  |  | Reference |
| --- | --- | --- | --- | --- | --- | --- | --- | --- | --- | --- |
|  |  |  | None (general population) | Children | Older adults | Persons with chronic medical conditions | Pregnant persons | Healthcare workers | Persons in congregate living settings |  |
| **Albania (UMIC)** | Pallas, 2020 | Cost-of-program |  |  |  |  |  | ? |  | [54] |
| **Argentina (UMIC)** | Giglio, 2012 | Cost-effectiveness (or cost-benefit, cost-utility) |  | ? |  |  |  |  |  | [58] |
| **Bangladesh (LIC)** | Bhuiyan, 2014 | Cost-of-illness | ? |  |  |  |  |  |  | [52] |
| **China (LMIC, UMIC)<sup>1</sup>** | Chen, 2019 | Cost-effectiveness (or cost-benefit, cost-utility) |  |  | ? |  |  |  |  | [60] |
|  | Gong, 2021 | Cost-of-illness | ? |  |  |  |  |  |  | [40] |
|  | Guo, 2012 | Cost-of-illness | ? |  |  |  |  |  |  | [47] |
|  | Jiang, 2020 | Cost-effectiveness (or cost-benefit, cost-utility) |  |  | ? |  |  |  |  | [62] |
|  | Lai, 2021 | Cost-of-illness |  | ? | ? | ? |  |  |  | [38] |
|  | Wang, 2013 | Cost-of-illness |  | ? |  |  |  |  |  | [31] |
|  | Wang, 2015 | Cost-of-illness |  | ? |  |  |  |  |  | [32] |
|  | Wang, 2019 | Cost-of-illness |  | ? |  |  |  |  |  | [37] |
|  | Wang, 2021 | Cost-of-illness |  | ? |  |  |  |  |  | [39] |
|  | Yan, 2021 | Cost-effectiveness (or cost-benefit, cost-utility) |  |  | ? |  |  |  |  | [64] |
|  | Yang, 2015 | Cost-of-illness | ? | ? | ? | ? |  |  |  | [33] |
|  | Yang, 2016 | Cost-of-program |  | ? | ? | ? | ? | ? |  | [55] |
|  | Yang, 2017 | Cost-of-illness | ? | ? | ? | ? |  |  |  | [35] |
|  |  |  | None<br>(general<br>population) | Children | Older<br>adults | Persons<br>with<br>chronic<br>medical<br>conditions | Pregnant<br>persons | Healthcare<br>workers | Persons in<br>congregate<br>living<br>settings |  |
|  | Yang, 2019 | Cost-effectiveness (or<br>cost-benefit, cost-utility) |  |  |  | ? |  |  |  | [61] |
|  | Yang, 2020 | Cost-effectiveness (or<br>cost-benefit, cost-utility) |  |  | ? |  |  |  |  | [63] |
|  | Yu, 2018 | Cost-of-illness |  | ? |  |  |  |  |  | [36] |
|  | Zhang, 2017 | Cost-of-illness |  | ? |  |  |  |  |  | [34] |
|  | Zhou, 2014 | Cost-effectiveness (or<br>cost-benefit, cost-utility) |  | ? |  |  |  |  |  | [59] |
| Colombia (UMIC) | Lara, 2018 | Cost-effectiveness (or<br>cost-benefit, cost-utility) |  | ? |  |  |  |  |  | [65] |
|  | Salcedo-Mejia,<br>2019 | Cost-of-illness |  | ? |  |  |  |  |  | [41] |
| El Salvador (LMIC) | Jara, 2019 | Cost-of-illness |  | ? |  |  |  |  |  | [43] |
| India (LMIC) | Koul, 2019 | Cost-of-illness |  |  |  | ? |  |  |  | [48] |
| Kazakhstan (UMIC) | Kovacs, 2014 | Cost-of-illness |  |  | ? |  |  |  |  | [42] |
| Kenya (LMIC) | Dawa, 2020 | Cost-effectiveness (or<br>cost-benefit, cost-utility) |  | ? |  |  |  |  |  | [78] |
|  | Emukule, 2019 | Cost-of-illness | ? | ? |  |  |  |  |  | [49] |
| Lao PDR (LMIC) | Ortega-Sanchez, 2021 | Cost-effectiveness (or<br>cost-benefit, cost-utility) |  |  | ? |  | ? | ? |  | [79] |
| Malawi (LIC) | Pecenka, 2017 | Cost-of-program |  |  |  |  | ? |  |  | [57] |
| Mali (LIC) | Orenstein, 2017 | Cost-of-illness AND<br>Cost-effectiveness (or<br>cost-benefit, cost-utility) |  |  |  |  | ? |  |  | [53] |
| Mexico (UMIC) | Betancourt- | Cost-effectiveness (or | ? |  |  |  |  |  |  | [67] |
|  |  |  | None (general population) | Children | Older adults | Persons with chronic medical conditions | Pregnant persons | Healthcare workers | Persons in congregate living settings |  |
|  | Craviato, 2021 | cost-benefit, cost-utility) |  |  |  |  |  |  |  |  |
|  | Falcon-Lezama, 2020 | Cost-effectiveness (or cost-benefit, cost-utility) |  | ? |  |  |  |  |  | [66] |
|  | Tapia-Conyer, 2021 | Cost-effectiveness (or cost-benefit, cost-utility) | ? | ? |  | ? |  |  |  | [68] |
| **Panama (UMIC)** | Jara, 2019 | Cost-of-illness |  | ? |  |  |  |  |  | [43] |
| **Romania (UMIC)** | Kovacs, 2014 | Cost-of-illness |  |  | ? |  |  |  |  | [42] |
| **South Africa (UMIC)** | Biggerstaff, 2019 | Cost-effectiveness (or cost-benefit, cost-utility) |  |  |  |  | ? |  |  | [70] |
|  | Edoka, 2021 | Cost-effectiveness (or cost-benefit, cost-utility) |  | ? | ? | ? | ? |  |  | [71] |
|  | Tempia, 2019 | Cost-of-illness | ? |  |  |  |  |  |  | [44] |
|  | Tempia, 2020 | Cost-of-illness |  | ? | ? | ? | ? |  |  | [45] |
|  | de Boer, 2018 | Cost-effectiveness (or cost-benefit, cost-utility) | ? |  |  |  |  |  |  | [69] |
| **Thailand (UMIC)** | Choosakulchart, 2013 | Cost-effectiveness (or cost-benefit, cost-utility) |  |  |  | ? |  |  |  | [72] |
|  | Kittikraisak, 2017 | Cost-effectiveness (or cost-benefit, cost-utility) |  | ? |  |  |  |  |  | [74] |
|  | Kittikraisak, 2018 | Cost-of-illness |  | ? |  |  |  |  |  | [46] |
|  | Meeyai, 2015 | Cost-effectiveness (or cost-benefit, cost-utility) |  | ? | ? |  |  |  |  | [73] |
|  | Riewpaiboon, 2021 | Cost-of-program |  |  |  |  | ? |  |  | [56] |
|  | Sribhutorn, 2018 | Cost-effectiveness (or cost-benefit, cost-utility) |  |  |  | ? |  |  |  | [75] |
|  |  |  | None<br>(general<br>population) | Children | Older<br>adults | Persons<br>with<br>chronic<br>medical<br>conditions | Pregnant<br>persons | Healthcare<br>workers | Persons in<br>congregate<br>living<br>settings |  |
|  | Suphanchaimat, 2020 | Cost-effectiveness (or cost-benefit, cost-utility) |  |  |  |  |  |  | ? | [76] |
| **Turkiye (UMIC)** | Akin, 2016 | Cost-effectiveness (or cost-benefit, cost-utility) |  |  |  | ? |  |  |  | [77] |
| **Ukraine (LMIC)** | Kovacs, 2014 | Cost-of-illness |  |  | ? |  |  |  |  | [42] |
|  | Kyi-Kokarieva, 2021 | Cost-effectiveness (or cost-benefit, cost-utility) |  |  |  |  |  | ? |  | [80] |
| **Vietnam (LMIC)** | Vo, 2017 | Cost-of-illness | ? | ? | ? |  |  |  |  | [50] |
|  | Vo, 2017 | Cost-of-illness | ? | ? | ? |  |  |  |  | [51] |
|  | de Boer, 2018 | Cost-effectiveness (or cost-benefit, cost-utility) | ? |  |  |  |  |  |  | [69] |

Abbreviations: LIC, low-income country; LMIC, lower-middle income country; SAGE, Strategic Advisory Committee of Experts on Immunization; UMIC, upper-middle income country; US\$, US Dollars
<sup>1</sup>China was classified as both UMIC and LMIC corresponding to studies before and after an upward change in World Bank classification in 2010.

**Figure S1:**
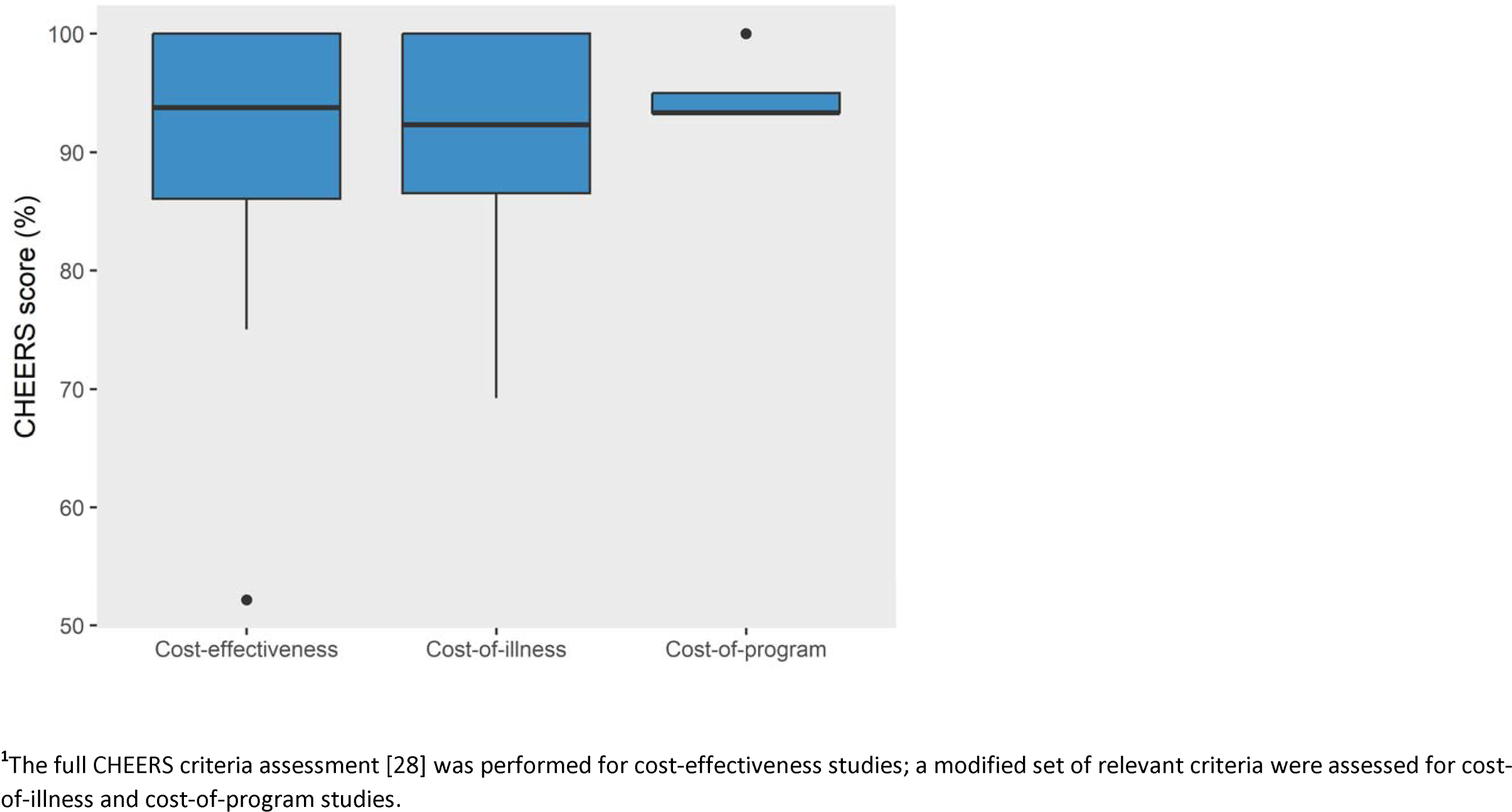
Distribution of modified Consolidated Health Economic Evaluation Reporting Standard (CHEERS) quality assessment scores.

**Figure S2:**
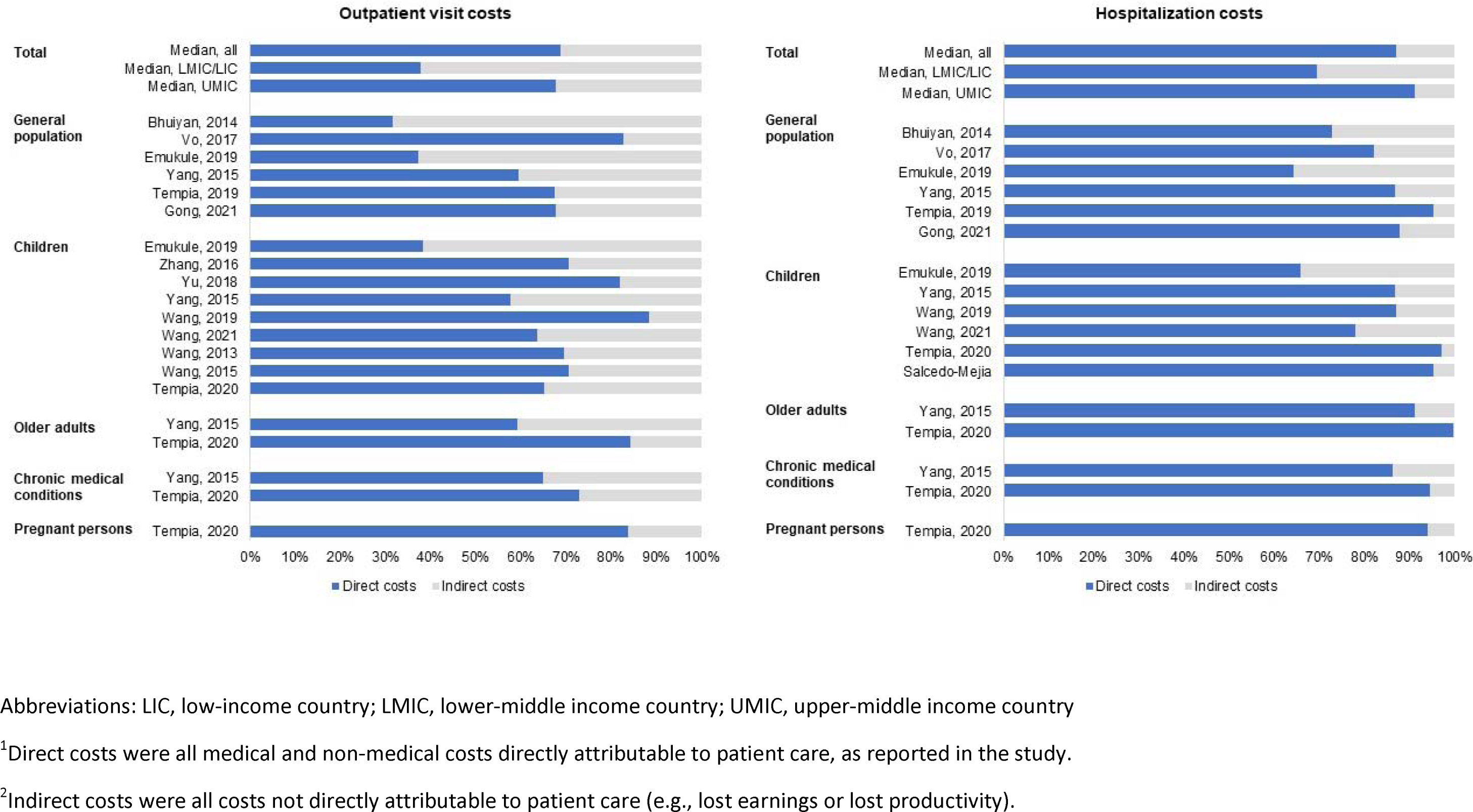
Contribution of direct and indirect costs to costs of influenza outpatient visits and hospitalizations, by Strategic Advisory Committee of Experts on Immunization (SAGE) target group, in low- and middle-income countries.

**Table S4:**
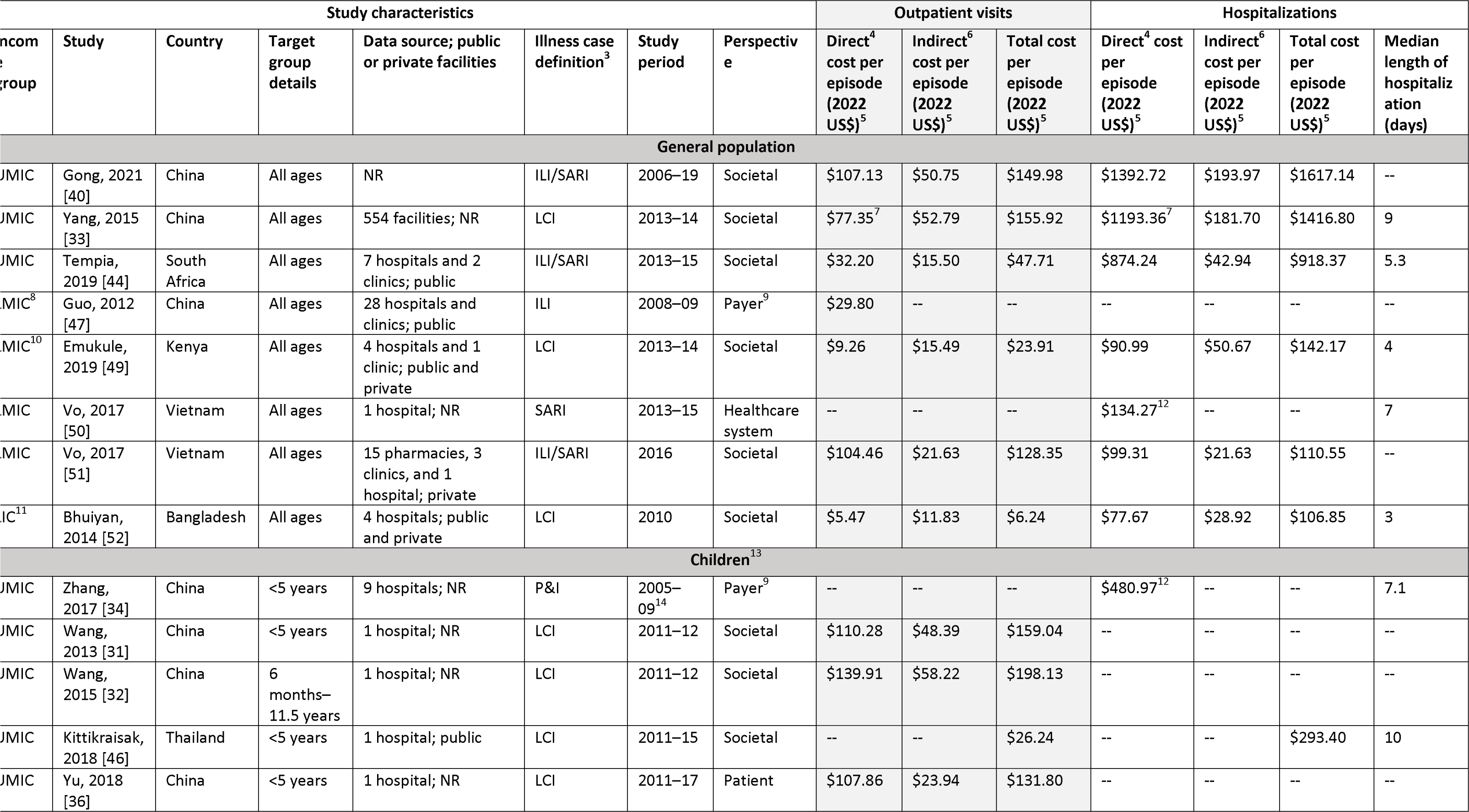
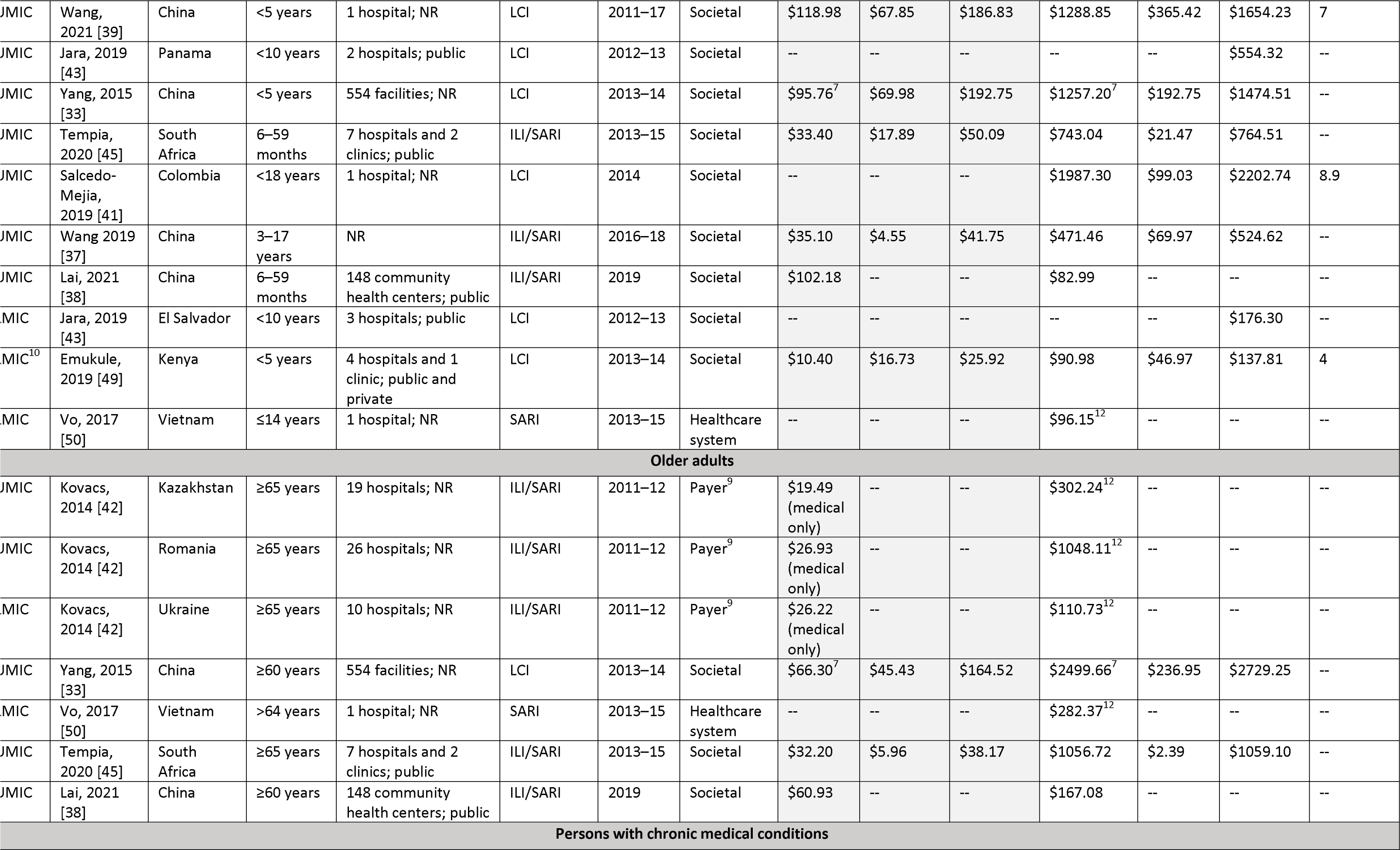
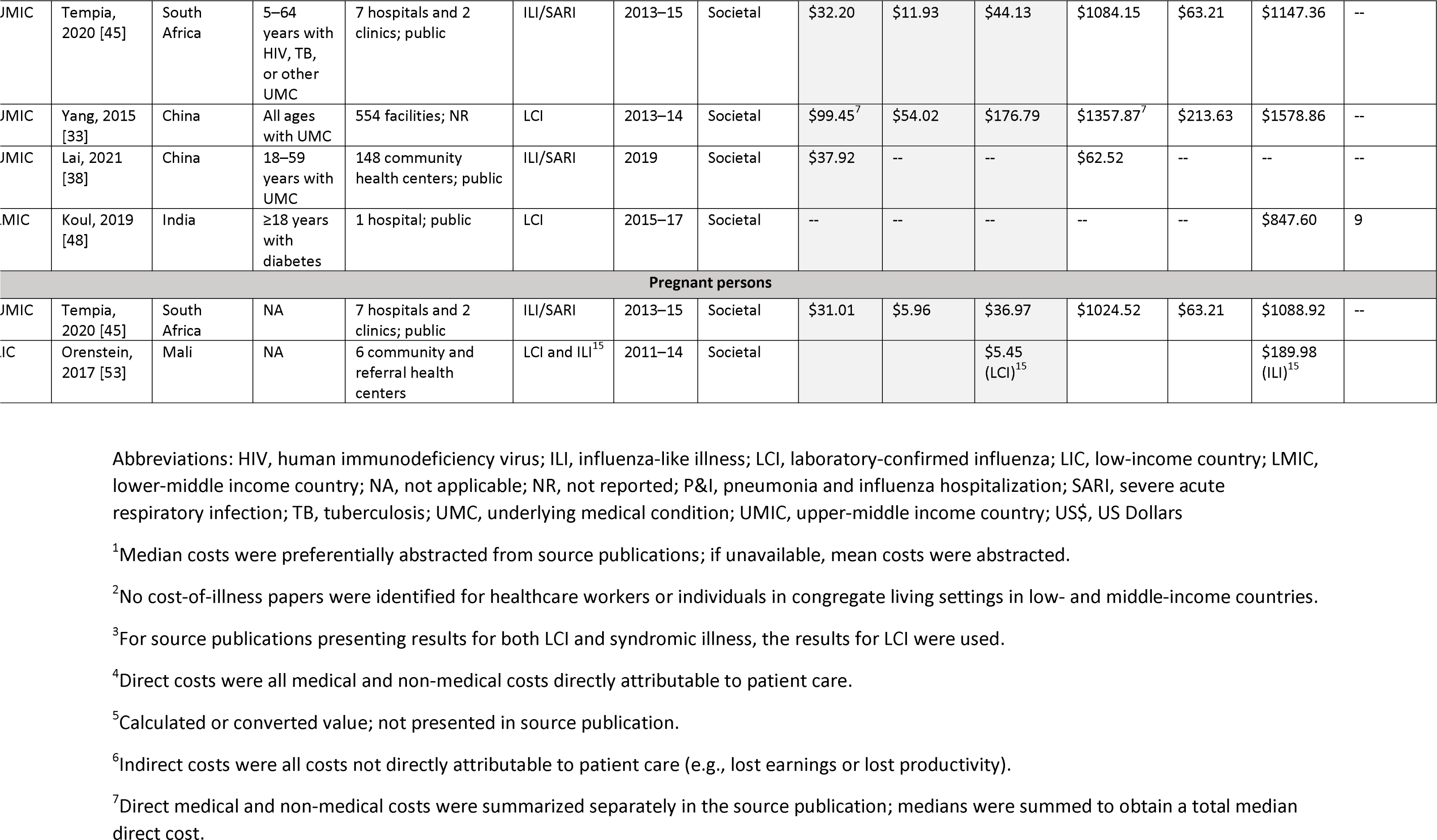
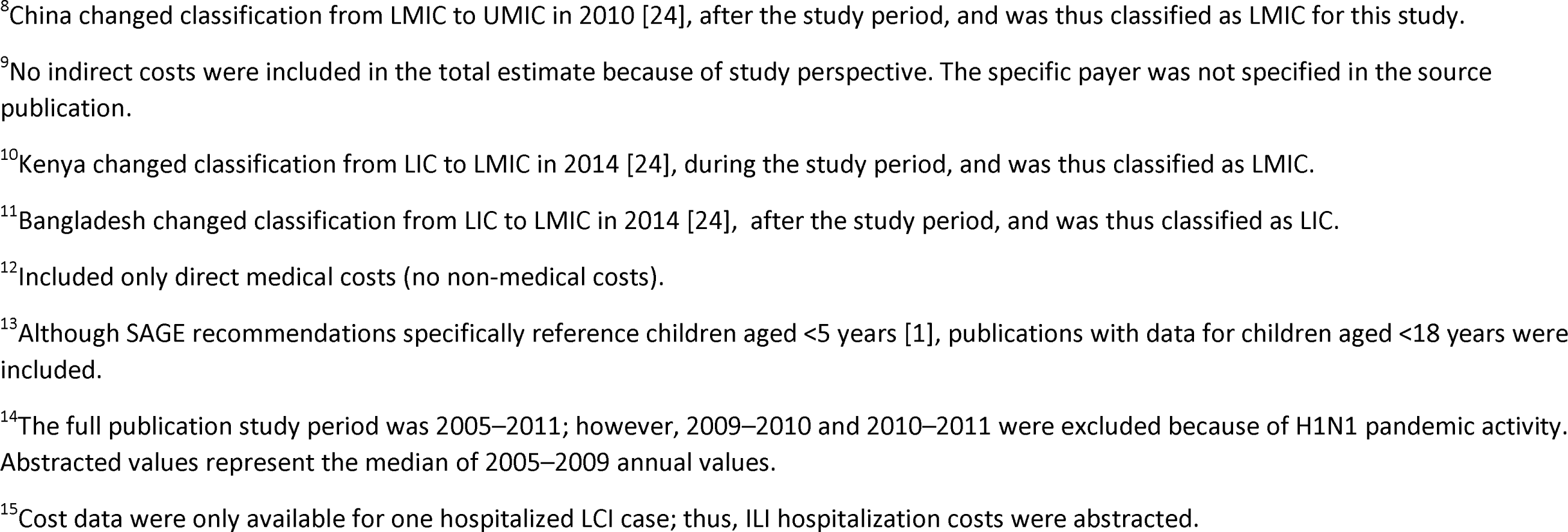
Costs of influenza illness^1^, by Strategic Advisory Committee of Experts on Immunization (SAGE) target group^2^ and disease severity (outpatient vs. hospitalized), in low- and middle-income countries.

**Table S5:**
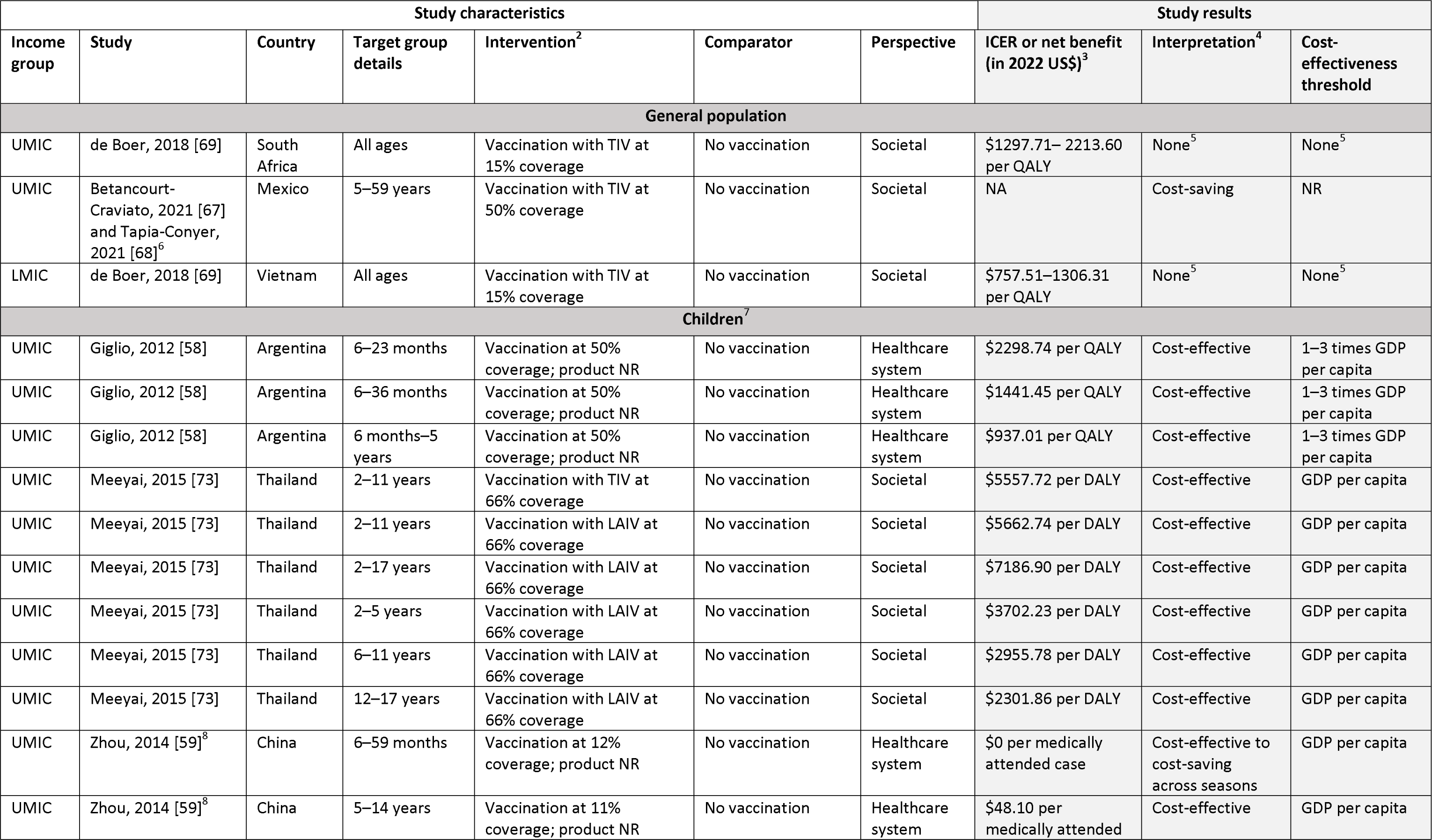
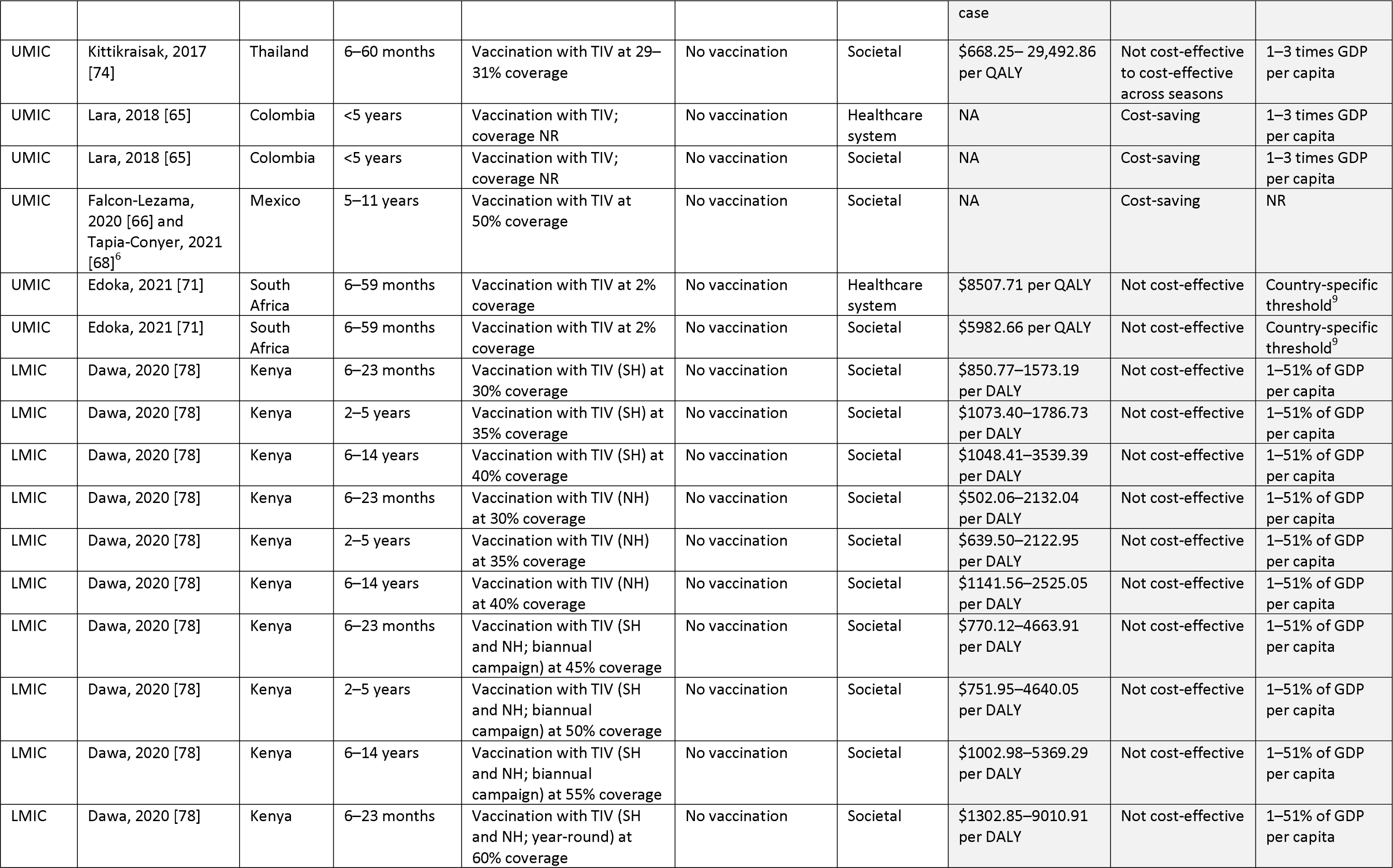
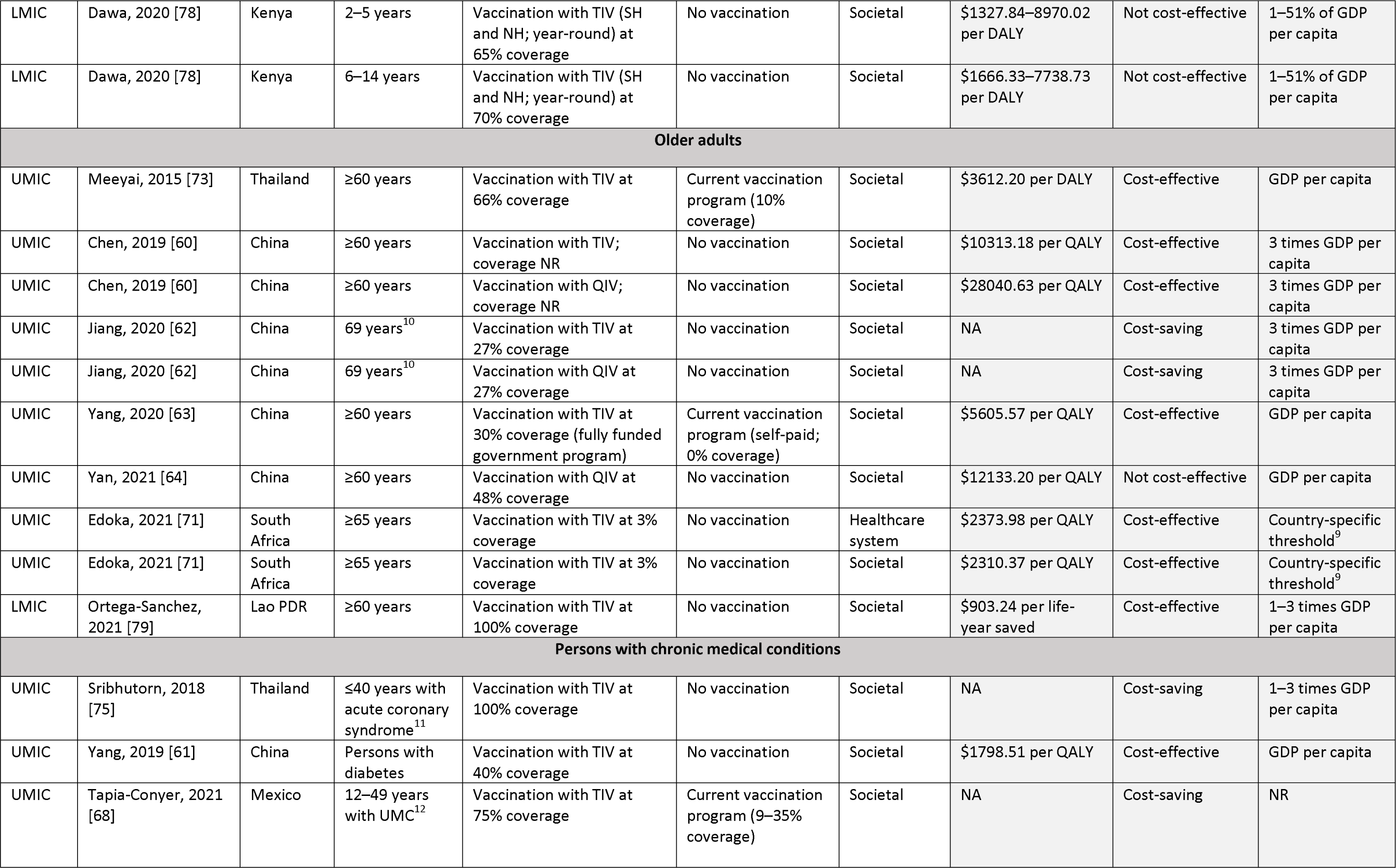
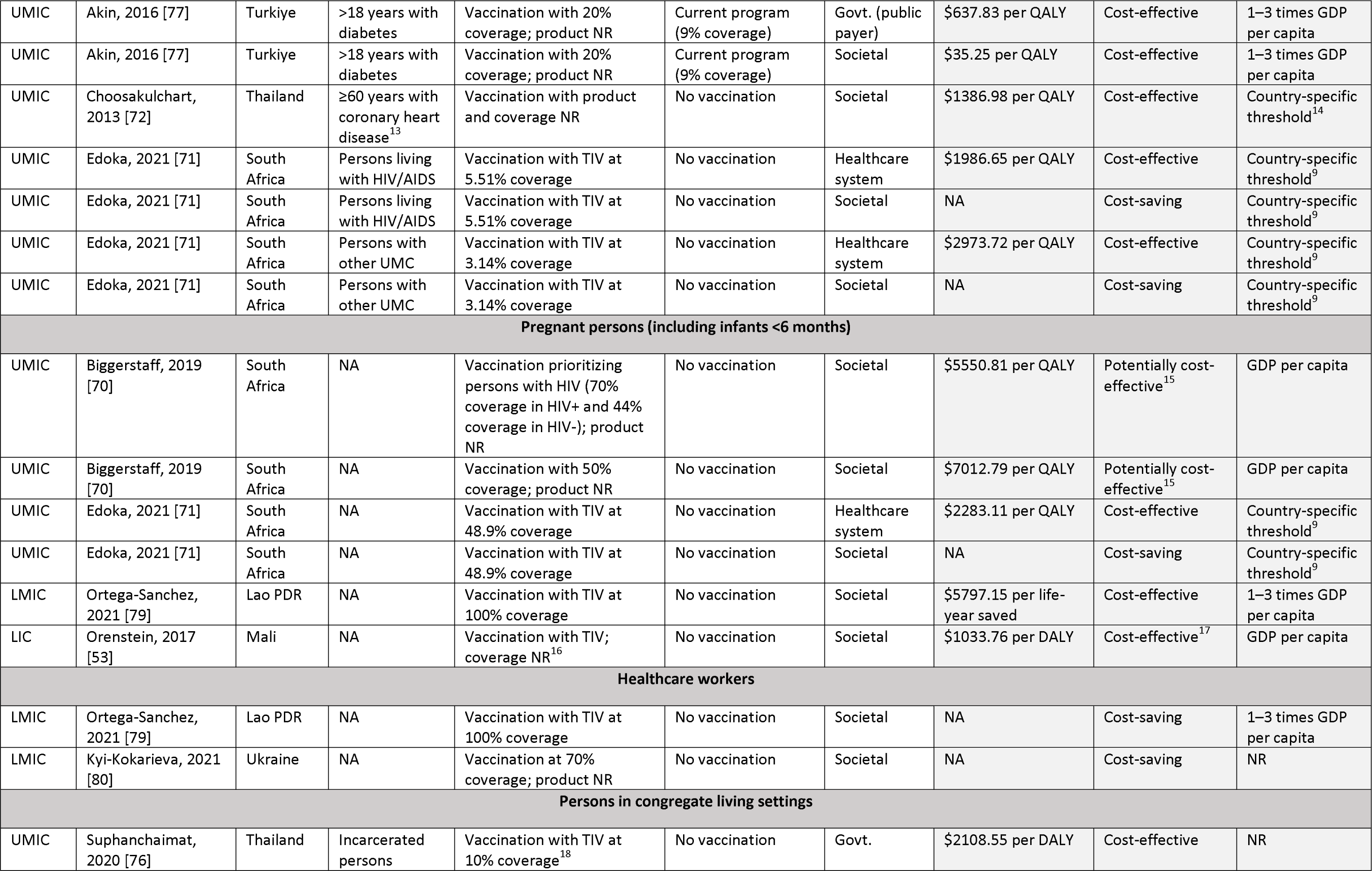
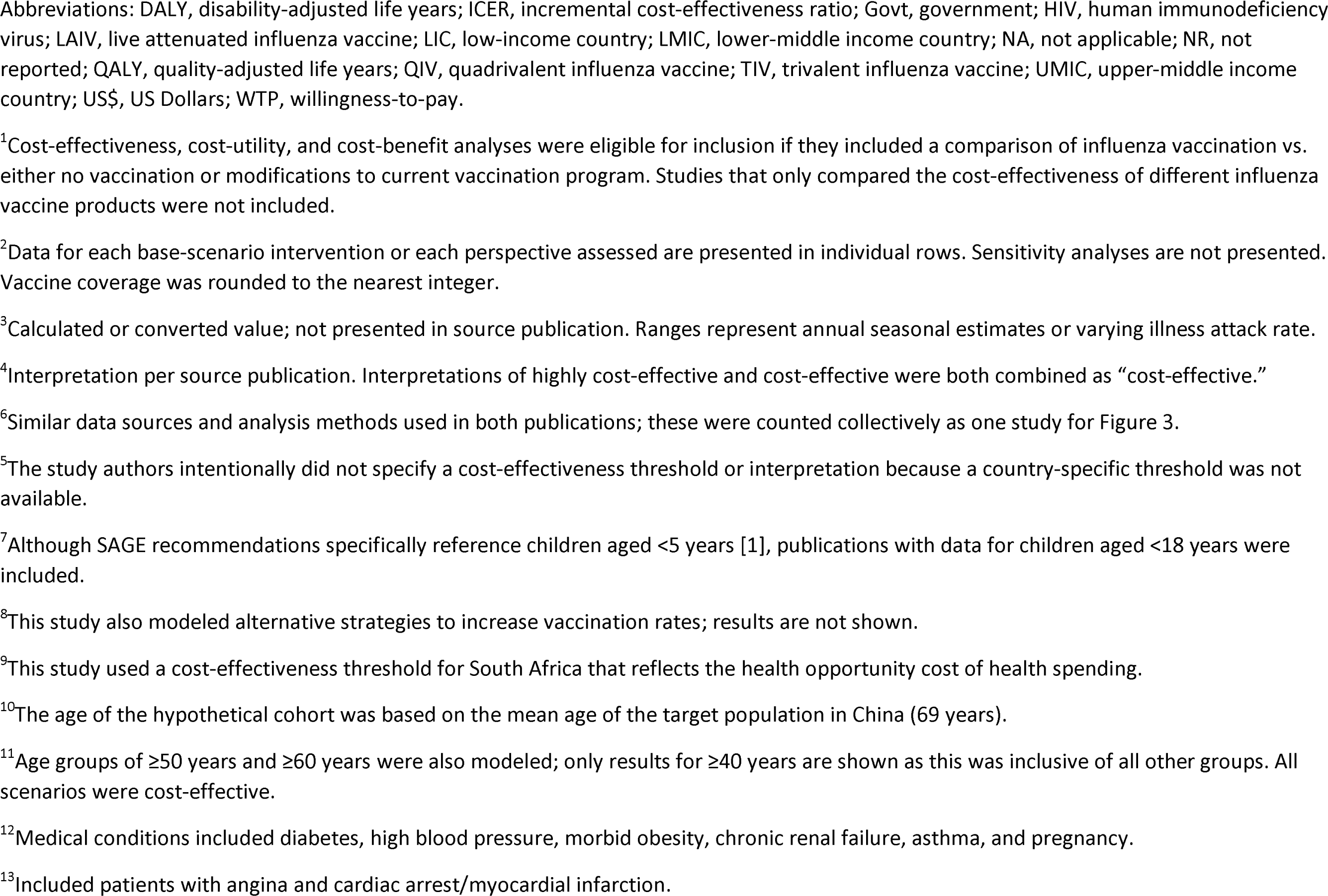
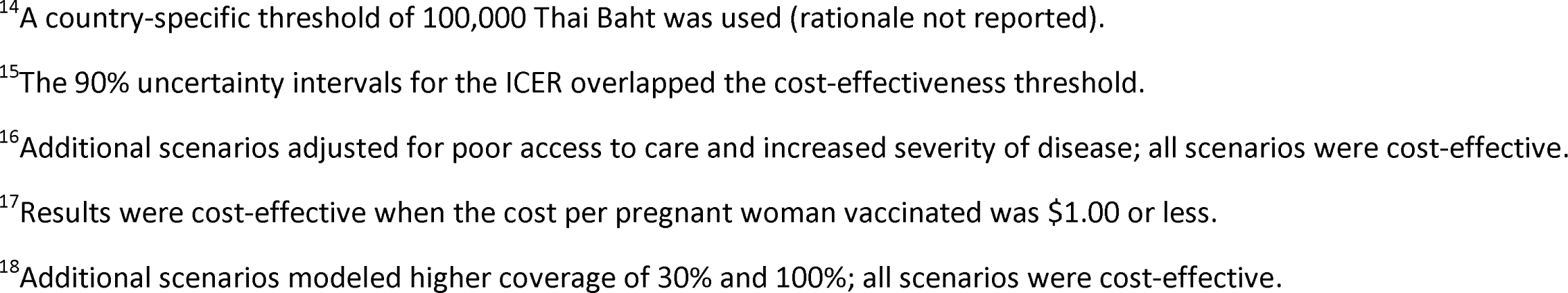
Cost-effectiveness of influenza vaccination^1^, by Strategic Advisory Committee of Experts on Immunization (SAGE) target group, in low- and middle-income countries.

